# Decisive Conditions for Strategic Vaccination against SARS-CoV-2

**DOI:** 10.1101/2021.03.05.21252962

**Authors:** Lucas Böttcher, Jan Nagler

## Abstract

While vaccines against SARS-CoV-2 are being administered, in most countries it may still take months until their supply can meet demand. The majority of available vaccines elicits strong immune responses when administered as prime-boost regimens. Since the immunological response to the first (“prime”) injection may provide already a substantial reduction in infectiousness and protection against severe disease, it may be more effective—under certain immunological and epidemiological conditions—to vaccinate as many people as possible with only one shot, instead of administering a person a second (“boost”) shot. Such a vaccination campaign may help to more effectively slow down the spread of SARS-CoV-2, reduce hospitalizations, and reduce fatalities, which is our objective. Yet, the conditions which make single-dose vaccination favorable over prime-boost administrations are not well understood. By combining epidemiological modeling, random sampling techniques, and decision tree learning, we find that single-dose vaccination is robustly favored over prime-boost vaccination campaigns, even for low single-dose efficacies. For realistic scenarios and assumptions for SARS-CoV-2, recent data on new variants included, we show that the difference between prime-boost and single-shot waning rates is the only discriminative threshold, falling in the narrow range of 0.01–0.02 day^−1^ below which single-dose vaccination should be considered.

## INTRODUCTION

After the initial identification of the novel severe acute respiratory syndrome coronavirus 2 (SARS-CoV-2) in Wuhan, China in December 2019, the virus quickly reached pandemic proportions and caused major public health and economic problems worldwide [1]. The disease associated with SARS-CoV-2 infections was termed coronavirus disease 2019 (COVID-19). As of June 28, 2021, the number of confirmed COVID-19 cases exceeded 180 million and more than 3.9 million COVID-19 deaths in more than 219 countries were reported [2]. Large differences between excess deaths and reported COVID-19 deaths across different countries suggest that the actual death toll associated with COVID-19 is even higher [3].

With the start, continuation and resuming of vaccination campaigns against SARS-CoV-2 in many countries [4], millions of people will receive partial and full immunization in the next months. The mRNA vaccines BNT162b2 (BioNTech-Pfizer) and mRNA-1273 (Moderna) received emergency use approval in the US and EU. When administered as prime-boost regimen, these vaccines have a reported protective efficacy of 95% [5] and 94.1% [6], respectively. An effectiveness evaluation of the BNT162b2 BioNTech-Pfizer vaccine shows that it may offer about 50% protection against SARS-CoV-2 infections about 2–3 weeks after receiving the first shot [7]. The adenovirus-based vaccine ChAdOx1 (Oxford-AstraZeneca) is being used in the UK, EU, and other countries with a reported single-shot regimen efficacy between 62–79% [8, 9]. Vaccine effectiveness against symptomatic disease for B.1.1.7 (Alpha) and B.1.617.2 (Delta) variants are reported to be 88% (Alpha) and 80% (Delta) for prime-boost regimens [10], while estimates of the effectiveness for hospitalization suggest 92% for Alpha and 94% for Delta [11, 12].

Taken together, the majority of currently available SARS-CoV-2 vaccines elicits strong immune responses against all studied variants when administered as prime-boost regimens. Yet, given the current distribution and production constraints, it may take months until the production of COVID-19 vaccines can meet the actual *global* demand. Similar to vaccination campaigns in previous disease outbreaks, it may therefore be a favourable alternative to administer a single vaccination dose to twice as many people. In 2016, a single-dose vaccination campaign against cholera was implemented in Zambia because of the insufficient number of vaccination doses that were available to complete a standard two-dose campaign [13]. Other vaccines, like the oral cholera vaccines that require two doses, are highly effective after a single dose but their protection is short lived compared to that obtained with prime-boost vaccination [14, 15].

Despite the clear advantages of single-dose vaccination campaigns, such as faster immunization of a larger number of people and lower vaccine-distribution infrastructure requirements and costs, any deviation from the immunologically favorable double-dose protocol may negatively affect the level of vaccination-induced immunity. An analysis of blood samples from COVID-19 patients suggests that the T cell response plays an important role in the long-term defense against SARS-CoV-2 [16] since antibody concentrations were found to decay faster than those of T cells that respond to SARS-CoV-2 epitopes. Clinical trial results [17] on the COVID-19 vaccine BNT162b1 show that the vaccination-induced CD4^+^ and CD8^+^ T cell responses are significantly reduced if no boost shot was administered, indicating that boost doses are important for T-cell-mediated immunity against SARS-CoV-2. In the same study, antibody concentrations in patients who received prime-boost regimes were found to be about 5 to 20 times higher than those observed in patients who only received a single vaccination dose, highlighting the need for boosting. Similar observations were made for type-1 inactivated poliovirus vaccine (IPV), for which clinical trial results [18] suggest that boost injections are needed to increase the level of neutralizing antibodies. However, for type-2 and 3 IPV, the first vaccination dose already elicits a neutralizing antibody response. In addition, single-dose vaccination may provide already a substantial degree of protection against infection, as confirmed in studies for BNT162b2 [7] and ChAdOx1 (Oxford-AstraZeneca) [19]. Yet, the mechanisms of vaccination-induced humoral (antibody-mediated) and cell-mediated immunity in SARS-CoV-2 is not well understood and data on immunity waning is scarce [20, 21].

Here, we study epidemiological population dynamics of SARS-CoV-2, where vaccine-induced protection levels, immunity waning, and other immunological factors are model parameters. Under which epidemiological conditions is single-dose vaccination favorable over prime-boost vaccination? This question is being controversially debated in many countries, including the US [22, 23], UK [24, 25], and Germany [26], as they are fearing the increasingly wide-spread of faster-spreading, more deadly SARS-CoV-2 mutants [27], such as the B.1.167.2 (Delta) variant, and the risk of collapsing health care systems [28].

The current controversy around prime and prime-boost vaccination strategies raises two connected questions, which we address in this paper: How do shortages in vaccine supplies and uncertainties in epidemiological parameters alter the possible advantage of single-dose over prime-boost vaccination? And how do possible differences in vaccine efficacy and loss of vaccine-induced immunity affect the decision boundary separating single-dose and prime-boost vaccination regimes in high-dimensional parameter space? By combining methods from epidemiological modeling, statistical mechanics [29, 30], and decision tree learning, we explore position, extent, and sensitivity of the decision boundary and provide a characterization of discriminative criteria [31], sufficiently simple and immediately accessible to decision makers.

## RESULTS

### Prime-first versus prime-boost vaccination

Different vaccination campaigns may lead to different proportions of infected, recovered, and deceased individuals at a given time. We study the differences between *prime-first* (Fig. 1) and prime-boost campaigns by accounting for a vaccination-induced reduction in transmissibility in a susceptible-exposed-infected-recovered-deceased (SEIRD)-based model [32] (see Materials and Methods and Fig. 1). To quantify the effect vaccination protocols have on the overall disease-induced fatality we use two fatality measures. The first measure is based on fatality rates, and the second one is based on cumulative deaths. Specifically, let *d*_1_ (prime-first) and *d*_2_ (prime-boost) be the maximum (daily) changes in the total number of deaths within the time horizon of about 10 months (*T* = 300 days). As a measure of the relative difference between *d*_1_ and *d*_2_, we use the relative fatality change (RFC-*δ*),

**FIG. 1.**
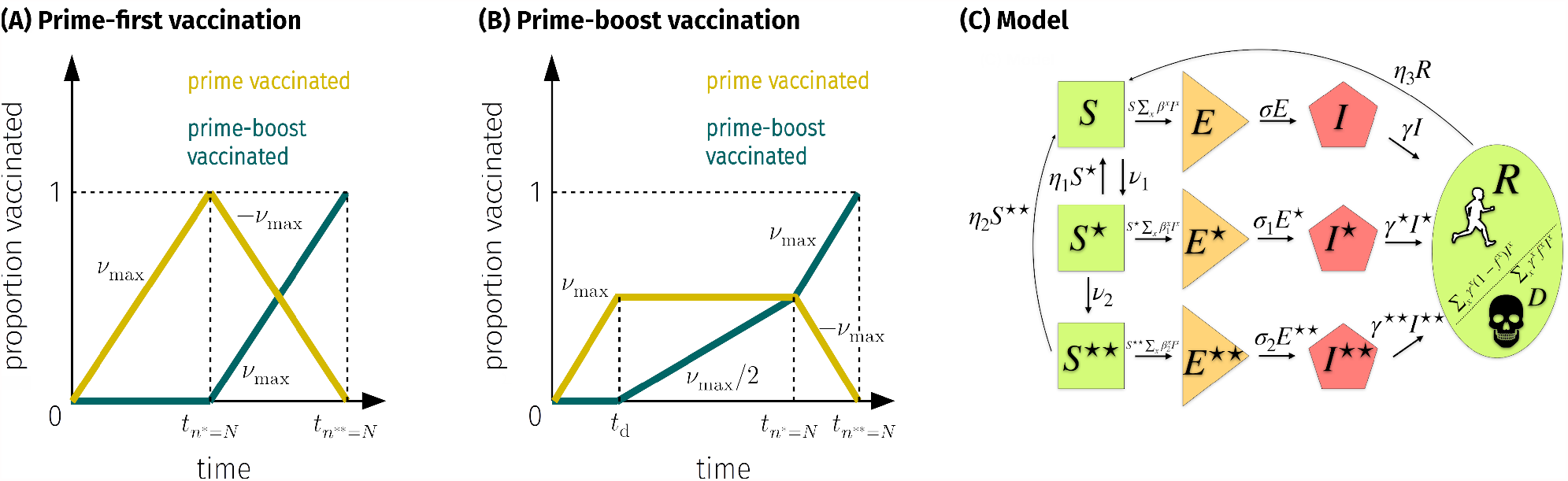
Schematic of vaccination campaigns and model. (A,B) Evolution of the number of individuals that received prime *(n*^★^) and prime-boost (*n*^★★^) shots in a population of size *N*. (A) Prime-first (★): Prime shots are administered with maximal rate as long as unvaccinated susceptible individuals exist. Then, boost shots are administered to prime-vaccinated individuals. (B) Prime-boost (★★) balances prime and boost shots equally. Boosting starts after *t*_*d*_ days. (C) In our model, susceptible individuals (*S, S*^★^, *S*^★★^) become exposed (*E, E*^★^, *E*^★★^) at rates *β, β*^★^, *β*^★★^ and transition to an infected state (*I, I*^★^, *I*^★★^) at rates *σ, σ*_1_, *σ*_2_. Infectious individuals either recover (*R*) or die (*D*) at rates ∑_*x*_(1 − *f*^*x*^)*γ*^*x*^ and ∑*_x_f*^*x*^*γ*^*x*^, respectively. Prime vaccination doses (★) are administered to susceptible individuals at rate *ν*_1_ and prime-vaccinated individuals receive a boost shot (★★) at rate *ν*_2_. Transitions from *S*^★^, *S*^★★^, and *R* to *S* (“waning immunity”) occur at rates *η*_1_, *η*_2_, and *η*_3_, respectively. Notation: ∑_*x*_ *β* ^*x*^ *I* ^*x*^= *βI* + *β* ^★^ *I*^★^ + *β* ^★★^*I*^★★^.

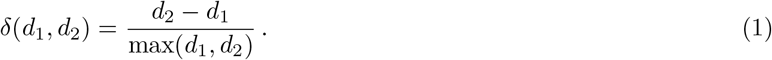

As a cumulative measure, we study the relative change in the cumulative number of deaths (RFC-Δ),

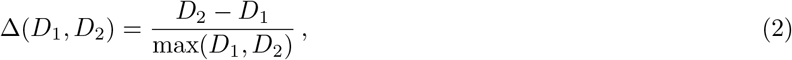

defined within the same time horizon as RFC-*δ*.

For both measures (1) and (2), a positive sign indicates more fatalities for prime-boost vaccination than for prime-first, while a negative sign indicates to favor prime-boost over prime-first campaigns. In the Materials and Methods, we show that and how the measures are correlated.

Current vaccination campaigns prioritize health care workers and vulnerable groups (e.g., elderly people with comorbidities) with a high risk of infection, leading to variations in vaccination rates. Further heterogeneity in model parameters may arise from infection rates that differ between age groups because of different degrees of susceptibility to infection [33] and different mobility characteristics. Our model accounts for these variations in epidemiological parameters through a large degree of parameterization. Nine different infection rates describe contacts between (susceptible and infectious) unvaccinated, single-dose vaccinated, and prime-boost vaccinated individuals. This large degree of parameterization can effectively account for possible correlations between age-group, transmissbility, and mobility. We therefore choose not to incorporate demographic compartmentalization in our model [30]. Yet, we study the effects of age-stratification, natural immunity waning, and effects from parameter constraints in separate scenarios.

### Vaccination-campaign-preference diagrams

To provide mechanistic insight into the population-level differences between prime and prime-boost vaccination campaigns, we study how RFC-*δ* and RFC-Δ are impacted by epidemiological parameters and epidemic state. As a function of two parameters, green domains as shown in Fig. 2 indicate excess deaths for prime-boost, while prime-boost is favored in red regions. The parameter ranges follow existing literature, or are chosen sufficiently broad to cover uncertainties. Empirical data [34] suggests an estimated range of the basic reproduction number *R*_0_ ∈ [1, 4] for the wild-type virus strain. Variants may be outside this range, in particular B.1.167.2 (Delta). Yet, in virtually all scenarios, the campaign preference does not change for larger values of *R*_0_ (see Materials and Methods for additional analyses). Differences in the waning rates *η*_1_ and *η*_2_ are not known at the present time, not even conclusive estimates [35], while clinical trials are still ongoing. Thus, we sample a broad parameter range, *η*_1_ − *η*_2_ ∈ [10^−4^, 10^−1^] day^−1^ with *η*_2_ = 3 × 10^−3^ day^−1^, which includes waning time-scales that were reported earlier for SARS-CoV [20]. For the initial infection disease prevalence, we assume the range *I*(0) ∈ [10^−4^, 10^−1^]. This range includes up to 10% infected individuals but may lie outside estimates of some places with a very high prevalence such as Manaus, Brazil [36] as faced in August 2020, and earlier estimates from New York City, USA [37]. For the range of the maximum vaccination rate *ν*_max_ we use [0, 10^−1^] day^−1^, which we inferred from current vaccination-campaign data [4].

**FIG. 2.**
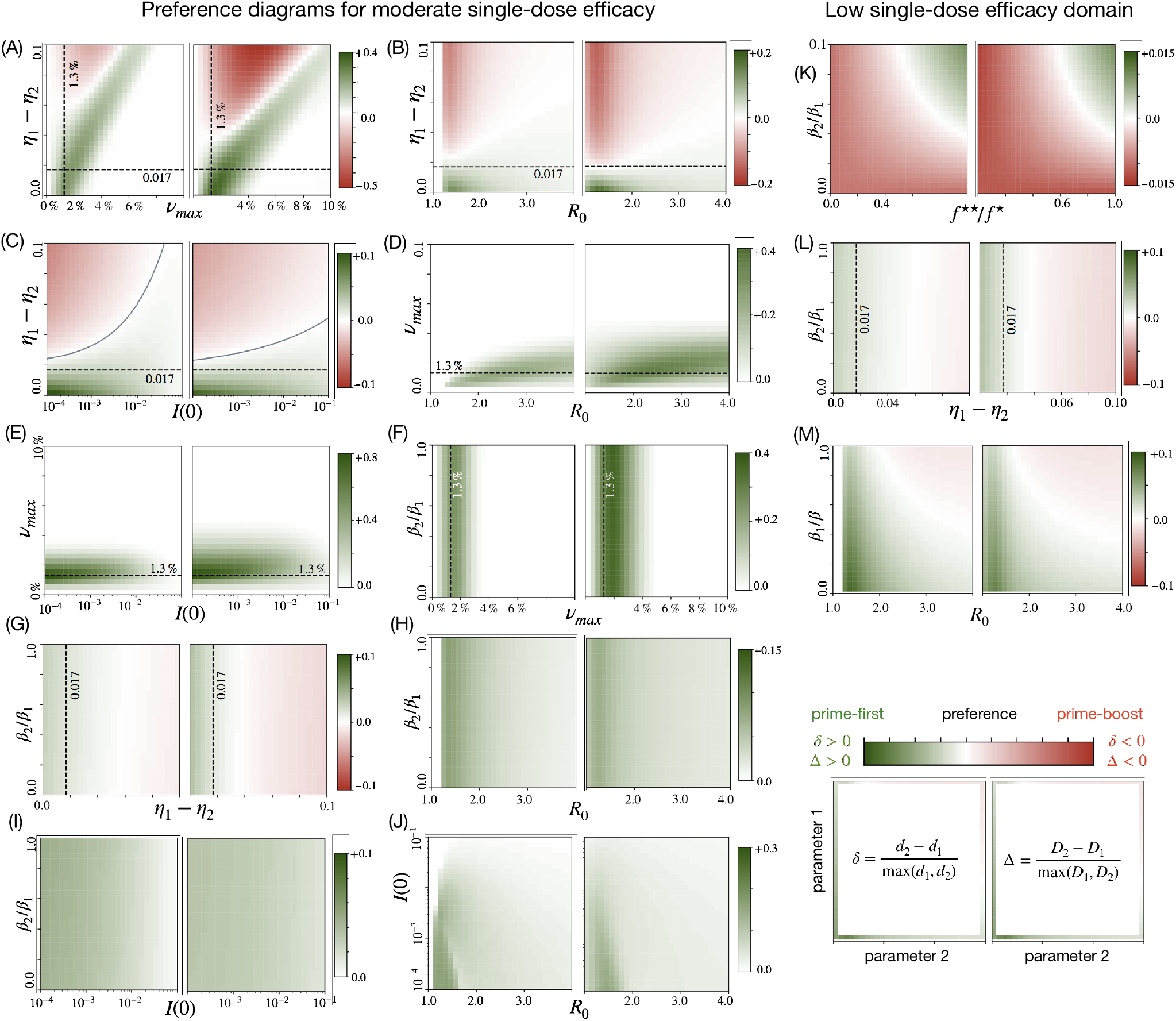
Vaccination-campaign-preference diagrams. For combinations of basic reproduction number *R*_0_, waning rate difference *η*_1_ − *η*_2_, initial disease prevalence *I*(0), maximum vaccination rate *ν*_max_, and relative efficacy for prime-first immunization (RE), *β*_2_ */β*_1_, we plot RFC-*δ* [Eq. (1)], and RFC-Δ [Eq. (2)]. Green-shaded regions indicate preference for prime (RFC-*δ >* 0, RFC-Δ > 0), red-shaded regions indicate preference for prime-boost (RFC-*δ <* 0, RFC-Δ < 0). (A-L): Parameter domain as in Tab. I, assuming a *moderate* single-dose efficacy, i.e. *β*_1_ = *β/*2, *f*^★^ = *f/*10. (K-M): *Low* single-dose efficacy domain: we set *β*_1_ = 0.9*β, f*^★^ = 0.6 × 10^−2^ = 0.6*f*, and all remaining parameters as in Tab. I. For (M) we varied *β*_1_ */β* and *β* (hence *R*_0_) and set all other parameters according to Tab. I. The ratios 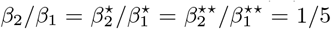 are also as in Tab. I. Dashed lines: Decisive threshold *η*_1_ − *η*_2_ = 0.017 day^−1^, and Israel’s vaccination rate as of Feb. 1, 2021 [4] (*ν*_max_ = 0.013 day^−1^). Solid line in (C): decision boundary as guide to the eye between *y* = *η*_1_ − *η*_2_ and *x* = *I*(0) as given by the following nonlinear relations: *y* = 0.4*x*^1*/*2^ + 0.02 (RFC-*δ*), and *y* = 0.06*x*^1*/*4^ + 0.017 (RFC-Δ).

We assume that the transmission rates *β*_1_ and *β*_2_ are proportional to the vaccine efficacies after single-dose and prime-boost vaccination, respectively. Thus, we identify the relative efficacy for single-dose immunization (RE) with the ratio *β*_2_*/β*_1_. Values close to one are favorable for prime-first campaigns, while a low RE disfavors prime-first.

In order to analyze the effect of RE on the effectiveness of prime and prime-boost vaccination campaigns, we study *β*_2_*/β*_1_ ∈ [10^−4^, 1] day^−1^. We choose this rather broad range to account for the lack of reliable data, in particular regarding new variants of SARS-CoV-2 and possible adverse effects in vaccine protection [38]. Parameters that are held constant in our simulations are listed in Tab. I (see Materials and Methods).

**TABLE I.** Overview of model parameters for scenarios without natural immunity waning. The listed parameter values are used when the associated model parameters are held constant in the parameter-space plots that we show in the results section. As initial fractions of infected and susceptible individuals, we use *I*(0) = 10^−2^ and *S*(0) = 1 − *I*(0).

| Parameter | Symbol | Value | Units | Comments/References |
| --- | --- | --- | --- | --- |
| infection rates $S$ | $\beta, \beta^*, \beta^{**}$ | $3/14, \beta/10, \beta/20$ | $[\text{day}^{-1}]$ | $\beta$ inferred from $R_0 = \beta/\gamma$ [34] |
| infection rates $S^*$ | $\beta_1, \beta_1^*, \beta_1^{**}$ | $\beta/2, \beta^*/2, \beta^{**}/2$ | $[\text{day}^{-1}]$ | estimate |
| infection rates $S^{**}$ | $\beta_2, \beta_2^*, \beta_2^{**}$ | $\beta/10, \beta^*/10, \beta^{**}/10$ | $[\text{day}^{-1}]$ | estimate |
| incubation rate $E$ | $\sigma$ | $1/5$ | $[\text{day}^{-1}]$ | [34, 58] |
| incubation rate $E^*$ | $\sigma_1$ | $1/5$ | $[\text{day}^{-1}]$ | estimate |
| incubation rate $E^{**}$ | $\sigma_2$ | $1/5$ | $[\text{day}^{-1}]$ | estimate |
| vaccination rate | $\nu_{\max}$ | $10^{-3}$ | $[\text{day}^{-1}]$ | [4] |
| waning rate (prime) | $\eta_1$ | $10^{-2}$ | $[\text{day}^{-1}]$ | estimate |
| waning rate (prime-boost) | $\eta_2$ | $3 \times 10^{-3}$ | $[\text{day}^{-1}]$ | estimate |
| waning rate (recovered) | $\eta_3$ | $0$ | $[\text{day}^{-1}]$ | estimate |
| resolution rate $I$ | $\gamma$ | $1/14$ | $[\text{day}^{-1}]$ | [52, 59] |
| resolution rate $I^*$ | $\gamma^*$ | $2\gamma$ | $[\text{day}^{-1}]$ | estimate |
| resolution rate $I^{**}$ | $\gamma^{**}$ | $4\gamma$ | $[\text{day}^{-1}]$ | estimate |
| fatality ratio $I$ | $f$ | $10^{-2}$ | - | [3, 60] |
| fatality ratio $I^*$ | $f^*$ | $10^{-3}$ | - | estimated from reported efficacy [56] |
| fatality ratio $I^{**}$ | $f^{**}$ | $10^{-3}$ | - | estimated from reported efficacy [56] |
| prime-boost delay | $t_d$ | $21$ | $[\text{day}]$ | [56] and Fig. 4 |

**TABLE II.** Sampling distributions with and without natural immunity waning. The listed parameter values and distributions are used in our random-sampling analysis. As initial fraction of susceptible individuals, we use *S*(0) = 1 − *I*(0) − *S*^★^(0) − *S*^★★^(0). We set *E*(0) = 0, *E*^★^(0) = 0, *E*^★★^(0) = 0, *I*^★^(0) = 0, *I*^★★^(0) = 0, *R*(0) = 0, *D*(0) = 0. The minimum and maximum values of *β* are *β*_min_ = *γ* and *β*_max_ = 4*γ*, respectively. A uniform distribution with boundaries *a* and *b* is indicated by 𝒰 (*a, b*).

| Parameter | Symbol | Value/Distribution | Units |
| --- | --- | --- | --- |
| infection rates $S$ | $\beta, \beta^*, \beta^{**}$ | $\mathcal{U}(\beta_{\min}, \beta_{\max}), \mathcal{U}(0, \beta), \mathcal{U}(0, \beta^*)$ | [day<sup>-1</sup>] |
| infection rates $S^*$ | $\beta_1, \beta_1^*, \beta_1^{**}$ | $\mathcal{U}(0, \beta), \mathcal{U}(0, \beta_1), \mathcal{U}(0, \beta_1^*)$ | [day<sup>-1</sup>] |
| infection rates $S^{**}$ | $\beta_2, \beta_2^*, \beta_2^{**}$ | $\mathcal{U}(0, \beta_1), \mathcal{U}(0, \beta_1^*), \mathcal{U}(0, \beta_1^{**})$ | [day<sup>-1</sup>] |
| incubation rate $E$ | $\sigma$ | $\mathcal{U}(0.2, 0.5)$ | [day<sup>-1</sup>] |
| incubation rate $E^*$ | $\sigma_1$ | $\mathcal{U}(0.2, 0.5)$ | [day<sup>-1</sup>] |
| incubation rate $E^{**}$ | $\sigma_2$ | $\mathcal{U}(0.2, 0.5)$ | [day<sup>-1</sup>] |
| vaccination rate | $\nu_{\max}$ | $\mathcal{U}(0, 0.02)$ | [day<sup>-1</sup>] |
| waning rate (prime) | $\eta_1$ | $\mathcal{U}(0, 0.1)$ | [day<sup>-1</sup>] |
| waning rate (prime-boost) | $\eta_2$ | $\mathcal{U}(0, \eta_1)$ | [day<sup>-1</sup>] |
| waning rate (recovered) | $\eta_3$ | 0 [scenarios w/o natural immunity waning]<br>$\mathcal{U}(0, 0.1)$ [scenarios w/ natural immunity waning] | [day<sup>-1</sup>] |
| resolution rate $I$ | $\gamma$ | 1/14 | [day<sup>-1</sup>] |
| resolution rate $I^*$ | $\gamma^*$ | $\mathcal{U}(\gamma, 2\gamma)$ | [day<sup>-1</sup>] |
| resolution rate $I^{**}$ | $\gamma^{**}$ | $\mathcal{U}(\gamma^*, 2\gamma^*)$ | [day<sup>-1</sup>] |
| fatality ratio $I$ | $f$ | $\mathcal{U}(10^{-3}, 10^{-1})$ | - |
| fatality ratio $I^*$ | $f^*$ | $\mathcal{U}(10^{-3}, f)$ | - |
| fatality ratio $I^{**}$ | $f^{**}$ | $f^*$ | - |
| prime-boost delay | $t_d$ | $\mathcal{U}(7, 35)$ | [day] |
| initially infected individuals | $I(0)$ | $\mathcal{U}(10^{-4}, 3 \times 10^{-1})$ | - |
| initially prime-vaccinated individuals | $S^*(0)$ | $\mathcal{U}(10^{-4}, 10^{-1})$ | - |
| initially prime-boost vaccinated individuals | $S^{**}(0)$ | $\mathcal{U}(10^{-4}, 10^{-1})$ | - |

The vaccination-campaign-preference diagrams (Fig. 2) suggest that prime vaccination campaigns are associated with a smaller death toll compared to prime-boost campaigns for a wide range of *R*_0_, maximum vaccination rates, epidemic states, and relative efficacy ratios (green-shaded regions in Fig. 2).

As the main result of our study, we identify a two-parameter threshold combination that separates vaccination-campaign preferences (dashed black lines in Fig. 2). For a sufficiently small waning-rate difference *η*_1_ − *η*_2_ ≲ 0.02 day^−1^ and a sufficiently low maximum vaccination rate *ν*_max_ ≲ 0.02 day^−1^, we observe that prime-first vaccination outperforms prime-boost vaccination in all projections where parameters are held constant as specified in Tab. I. In the projections involving *η*_2_ − *η*_1_, prime-boost preference is observed if immunity wanes significantly faster for prime-vaccinated individuals than for prime-boost vaccinated individuals.

All projections in Fig. 2 combined suggest that prime-boost vaccination should only be favored for *ν*_max_ ≳ 0.02 day^−1^, which largely exceeds SARS-CoV-2 immunization rates worldwide [4].

How a relatively low single-dose efficacy affects the preference for each campaign is shown in Fig. 2(K–M). In Fig. 2(K,L) we assume a transmission reduction of only 10% after single-dose immunization, *β*_1_ = 0.9*β*, together with a 40% reduction in fatality, *f*^★^ = 0.6 × 10^−2^ = 0.6*f*, and all other parameters as in Tab. I. This “low single-dose efficacy” domain is comparable with current estimates of vaccine effectiveness of BioNTech-Pfizer and Oxford-AstraZeneca against symptomatic disease for Alpha (49%) and Delta (31%) variants [10, 11]. Yet, it represents substantial less efficacious single-dose vaccine regimens than those for BioNTech-Pfizer or AstraZeneca against Alpha (78%) and Delta (75%) regarding hospitalization [11, 12].

In addition, the low single-dose efficacy domain is characterized by the occurrence of additional prime-boost preference regions in parameter space [red-shaded regions in Fig. 2(K,L)]. Yet, even if the fatality rates of prime-first and prime-boost deviate substantially, *f*^★★^*/f*^★^ ≲ 0.8, only for low values of the relative prime-first efficacy RE = *β*_2_*/β*_1_ ≲ 0.1, preference for prime-boost is observed [Fig. 2(K), shown range 0 ≤ *β*_2_*/β*_1_ ≤ 0.1]. Given this range and current data on SARS-CoV-2 [7, 11, 19], the diagram suggests preference for prime-first. Regarding the waning rate difference, *η*_1_ − *η*_2_, a low single dose-efficacy does not suggest a threshold lower than 0.017 day^−1^ for prime-first preference [Fig. 2(L)].

Figure 2(M) shows the dependence of RFC-*δ* and RFC-Δ on *β*_1_*/β* and *R*_0_. Values of *β*_1_*/β* ≈ 1 indicate a very low single-shot efficacy, whereas *β*_1_*/β* ≈0 indicates an unrealistically high efficacy. For large *R*_0_ and very low single-shot efficacies, *β*_1_*/β* ≳ 0.8, prime-boost is preferred over prime-first [Fig. 2(M)]. These parameters are, however, unlikely to be characteristic of SARS-CoV-2 [7, 19], recent data on the Delta variant included [11].

Finally, the waning-rate threshold *η*_1_ − *η*_2_ = 0.017 day^−1^ robustly separates prime-first and prime-boost preference regions for varying natural immunity waning rates and empirical vaccination time series data (see Materials and Methods). The waning-rate threshold below which preference for prime-first is observed depends only weakly on the initial infection prevalence: it slightly increases as *I*(0) decreases, *η*_1_ − *η*_2_ ≲ 0.010 − 0.017 day^−1^ for *I*(0) = 10^−5^ − 10^−2^ (see Materials and Methods).

The presented campaign preference diagrams are two-dimensional projections of a 25-dimensional parameter space, with the majority of parameters kept arbitrarily fixed (Tab. I). Hence, we examine next whether the preference for prime-first vaccination is supported by other independent methods.

### High-dimensional parameter space Monte Carlo sampling

Thus far, our results suggest a pronounced preference for prime-first vaccination for a wide range of key epidemiological parameters. To further substantiate this conclusion, we performed Monte Carlo sampling of the entire 25-dimensional parameter space (see Materials and Methods). For the analyzed high-dimensional parameter space, our results support that prime-boost-preference occurs significantly less frequently than samples indicating an advantage of prime-first vaccination.

The relative frequencies of samples for which prime-boost vaccination outperforms prime-first vaccination, characterized by RFC-*δ <* 0 and RFC-Δ < 0, are estimated as 7.9% [standard error (SE): 0.2%] and 23.2% (SE: 0.4%), see orange bars in Fig. 3(a). For waning rate differences *η*_1_ − *η*_2_ ≤ 0.056 day^−1^ and vaccination rates *ν*_max_ ≤ 0.047 day^−1^, we find that the proportions of prime-boost-preference samples are 7.0% (SE: 0.2%) for RFC-*δ <* 0 and 15.4% (SE: 0.3%) for RFC-Δ < 0 [beige bars in Fig. 3(a)]. Further restricting the parameter space using the condition *η*_1_ − *η*_2_ < 0.017 day^−1^ (dashed black lines in Fig. 2) and currently reported vaccination rates *ν*_max_ < 0.013 day^−1^ [4] leads to proportions of prime-boost-preference samples of 8.5% (SE: 0.2%) for RFC-*δ <* 0 and 6.9% (SE: 0.2%) for RFC-Δ < 0 [blue bars in Fig. 3(a)].

**FIG. 3.**
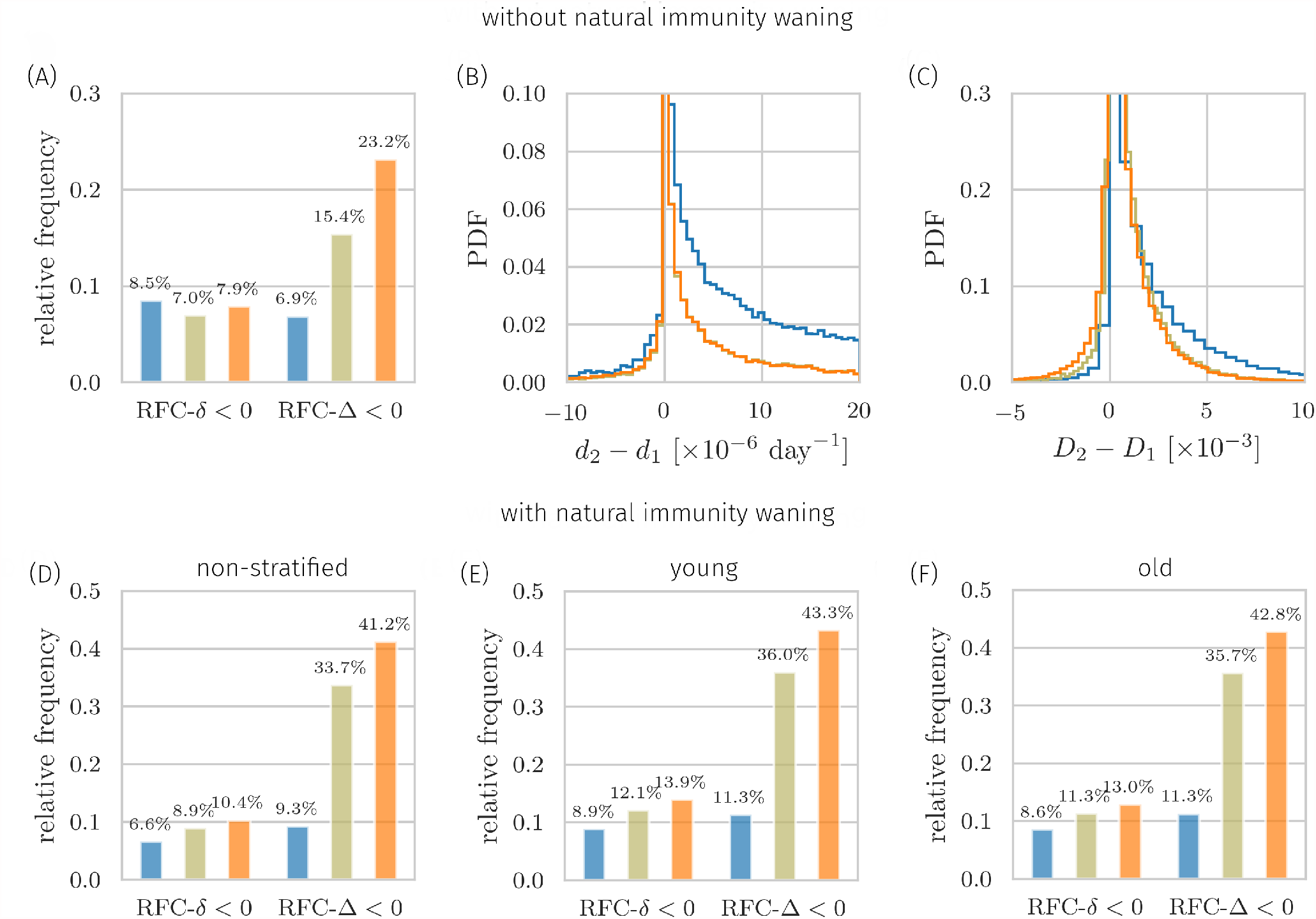
Monte Carlo sampling of entire and restricted high-dimensional parameter spaces. Unconditioned data (orange) is compared with two conditioned, (i) data shown in beige: *ν*_max_ ≤ 0.047 day^−1^ and *η*_1_ −*η*_2_ ≤ 0.056 day^−1^, and (ii) data shown in blue: *ν*_max_ ≤ 0.013 day^−1^ and *η*_1_ − *η*_2_ ≤ 0.017 day^−1^. Thresholds (i) are inferred from our decision tree analysis (Materials and Methods). (A) Relative frequency of prime-boost-preference samples (RFC-*δ <* 0, RFC-Δ < 0) for the three datasets. Error bars are below 0.2% (not shown). (B) Probability density function (PDF) of the difference between death rates *d*_2_ (prime-boost) and *d*_1_ (prime). For the conditioned data, averages of *d*_1_ −*d*_2_ are about 3 × 10^−5^ day^−1^ (blue curve) and about 5 × 10^−6^ day^−1^ (beige curve), larger than the mean 4 × 10^−6^ day^−1^ of the unconditioned data. (C) PDF of the difference between the total number of deaths *D*_2_ (prime-boost) and *D*_1_ (prime). For the conditioned data, the means of *D*_1_ −*D*_2_ are about 2 × 10^−3^ (blue curve) and about 9 × 10^−4^ (beige curve), larger than the mean 7 × 10^−4^ of the unconditioned data. The presented data are based on 5 × 10^4^ (blue and orange curves) and about 4.4 × 10^4^ (beige curves) samples of the entire dimensional parameter space. (D–F) Relative frequency of prime-boost-preference samples (RFC-*δ <* 0, RFC-Δ < 0) for additional datasets with the same constraints as used in (A). Error bars are below 0.5% (not shown). (D) We sampled natural immunity waning rates *η*_3_ from the distribution 𝒰 (0, 0.1) day^−1^. All remaining parameters are specified in Tab. II. (E) Fatality rates are *f* ∼ 𝒰 (10^−4^, 10^−3^), *f* ^∗^ ∼ 𝒰 (10^−4^, *f*), and *f* ^∗^ = *f* ^∗ ∗^. Natural immunity waning rate *η*_3_ is sampled from 𝒰 (0, 0.1) day^−1^. All remaining parameters are specified in Tab. II. (F) Fatality rates are *f* ∼ 𝒰 (10^−3^, 10^−2^), *f* ^∗^ ∼ 𝒰 (10^−3^, *f*), and *f* ^∗^ = *f* ^∗ ∗^. Natural immunity waning rate *η*_3_ is sampled from 𝒰 (0, 0.1) day^−1^. All remaining parameters are specified in Tab. II.

This means that constraining the studied parameter space by lowering *ν*_max_ and *η*_1_ − *η*_2_ results in a substantially enhanced preference for prime-first in terms of reduced excess deaths, RFC-Δ. In contrast, we find that the proportion of prime-boost preference samples is almost unaffected by the chosen parameter restrictions, which is indicated by the observed narrow rage between 7 and 9% [Fig. 3(a)]. This supports the robustness of our results. Independent of the threshold combination, for randomly sampled parameters, prime-first is robustly preferred regarding RFC-*δ*. In addition, domination of prime-first preference is observed in the projections for two-parameter combinations (Fig. 2).

The discriminative power of the thresholds is also supported by random sampling results for different risk groups and in situations with natural immunity waning [Fig. 3(D–F)]. For further details, see Materials and Methods.

### Decision tree learning

As another independent method for determining decisive conditions for strategic vaccination campaigns, we performed binary decision tree learning with repeated stratified cross validation [39, 40]. This technique has proven useful to extract the most discriminative features in high-dimensional data. Our analysis suggests that *ν*_max_ and *η*_1_ − *η*_2_ are the most discriminative parameters within the 25-dimensional parameter space (see Materials and Methods). For the samples that we generated according to the distributions listed in Tab. I (orange lines and markers in Fig. 3), and the constraints *ν*_max_ ≤ 0.047 day^−1^ and *η*_1_ − *η*_2_ ≤ 0.056 day^−1^, about 70% of vaccination preferences of simulated scenarios are correctly predicted (see Materials and Methods for accuracy scores and details). Additionally constraining the parameter space with the thresholds that we used in the previous paragraph (beige and blue lines and markers in Fig. 3), results in prime-first preference for 93% of the parameter space volume. This suggests that for realistic vaccination rates, the vaccination-dose-dependent immunity waning rate difference is the only highly discriminative factor.

## DISCUSSION

Effective vaccination protocols are crucial to achieve a high immunization coverage, especially if vaccination supplies are limited. The ongoing debate on the most effective way of distributing prime-boost regimens against SARS-CoV-2 has been sparked by arguments suggesting that, from an epidemiological perspective, single-dose vaccination protocols may be more effective than immediate prime-boost administration given the current supply shortages [22, 24, 26, 41, 42]. For many COVID-19 vaccines, prime-boost protocols are considered immunologically efficient due to their ability to elicit strong and long-term humoral and cellular immune responses [17]. Immunologically efficient vaccination protocols, however, may be not epidemiologically favorable, in particular for exponentially increasing infection numbers and vaccine doses shortages on times scales of months. We have studied the effect of relevant immunological and epidemiological parameters (e.g., vaccine efficacy and immunity waning) on a possible advantage of prime-first over prime-boost vaccination by combining epidemiological modeling, methods from statistical mechanics, and decision tree learning. We have identified and studied decision boundaries separating the parameter regimes in which one or the other vaccination protocol is preferable. Our results suggest that prime-first campaigns are associated with a lower death toll compared to prime-boost vaccination campaigns, even for relatively high vaccination rates, and more surprisingly, for low single-dose efficacies, which is in contrast to existing literature [7, 19, 41, 43].

A related study [41] compares single-dose and prime-boost vaccination campaigns against SARS-CoV-2, without accounting for immunity waning. This study reports that single-dose vaccination campaigns make optimal use of resources in the short term, given a sufficiently large single-dose efficacy that they identify as the main discriminative factor. In contrast, our study calls attention to immunity waning and the vaccination rate as the highly discriminative factors, while we find that vaccine efficacies are less discriminative.

Previous works consistently emphasize that due to the complexity of underlying models and limitations from available data, a vaccine campaign recommendation can only be given, once the precision in all key epidemiological parameters becomes sufficiently high. Laubenbacher *et al*. [44] highlight the need of further data collection and model integration in infectious disease modeling, which are important steps to better estimate immunity waning rates and vaccine effectiveness [7, 14, 43].

Saad-Roy *et al*. [45] focus on the long-term effects of waning and evolutionary immune response in a highly parameterized model. Certain scenarios they analyze suggest that single-dose campaigns may be favorable for some time scales but not for others, depending on a combination of parameters, waning rates included. In contrast, for the critical time scale of months, we provide a preference criterion based on the waning rate difference as the only discriminative threshold.

Preference for prime-first vaccination is not unexpected. For the initial inter-dose interval time, both vaccination strategies are identical since, regardless of the chosen strategy, booster jabs are not yet administered. In the subsequent time interval twice as many susceptible individuals can be immunized with a prime-first protocol compared to prime-boost vaccination. This means, about 50% of individuals who could have received a shot will actually remain unvaccinated. Let us refer to this unvaccinated group as *group A* and denote with *group B* those that receive both shots in the prime-boost campaign. One can assume that the infection rates of individuals in group *A* are larger than those of individuals in group *B*, who benefit from a more effective immune response. As a result, higher transmission in group *A* is the expected dominating differential adverse effect. As expected for effective prime-boost vaccines, one may assume that the prime-boost infection rate, *β*_2_, and the fatality rate, *f*^★★^, are significantly lower than their counterparts for prime-first. Thus, the effective transmission rate for group *A* and *B* combined is dominated by group *A*’s rate but not critically dependent on *β*_2_, which intuitively explains why the relative efficacy ratio RE = *β*_2_*/β*_1_ is not a highly discriminative factor.

For very low single-shot efficacies, or very high single-shot disease-induced fatality rates, the single-dose efficacy *β*_1_*/β*, the relative prime-first efficacy RE, and *R*_0_, may be discriminative, depending on the circumstances. Current data on SARS-CoV-2 vaccination campaigns [7, 19], however, suggest that those parameter combinations are unlikely to occur. Furthermore, if immunity wanes substantially faster after the first shot than after the additional booster jab, prime-boost vaccination may become favorable over prime-first, depending on *R*_0_. Unvaccinated and susceptible individuals should also receive both vaccination shots if a few percent of a jurisdiction’s total population can be vaccinated daily. However, even for the relatively large vaccination rate of about ∼ 1% per day, as realized in Israel [4], our analyses suggest that prime-first vaccination is still favorable over prime-boost campaigns.

A recent study in single-dose vaccinated SARS-CoV-2 patients infected with B.1.351 (Beta) or B.1.617.2 (Delta) variants showed neutralizing antibody concentrations below the quantitative limit of detection [46]. Does this well recognized study challenge our findings? While antibody titres correlate with protection against severe disease [47], they are only a single component of the intricate immune response and are not a necessary condition for effectiveness of vaccines against symptomatic disease or hospitalization. In fact, recent effectiveness estimates suggest that the first dose of BNT162b2 and ChAdOx1 is about 75% effective against hospitalization after an infection with the Delta variant (78% for Alpha, B.1.1.7) [11, 12]. These data strongly support our conclusions, given the high correlation between hospitalization and fatality.

But what about data on vaccine effectiveness against symptomatic disease? The effectiveness estimates for Alpha and Delta variants as reported in [11, 12] are as low as 31% for single-dose immunization, compared to 80% for two doses. While efficacies below 50% may strongly suggest that the population should get as soon as possible both immunization shots and not only one, our study finds that prime-first vaccination should be considered if the primary health objective is minimizing hospitalizations and fatalities. This means that vaccination campaigns that deviate from the recommended immunization protocol are particularly relevant in countries facing a possible health crises from emerging variants such as the Delta variant [48–50].

In summary, our results contrast existing literature [14, 41, 43, 45] in the sense that not all key epidemiological data are required to be collected to identify most effective vaccination protocols. Instead, our analysis suggests that even for a large degree of uncertainty in key epidemiological data, prime-first vaccination is robustly preferred over prime-boost vaccination—if the waning rate difference between prime-first and prime-boost is sufficiently small. For realistic scenarios specific to SARS-CoV-2, we found this threshold to be in the narrow range of 0.01–0.02 day^−1^. Unfortunately, to date, there is no reliable data available on waning time scales [20, 35], although recent estimates may suggest that no significant waning occurs for ChAdOx1 (Oxford-AstraZeneca) in the first 90 days after receiving the first shot [19]. Yet, once vaccination-dependent waning rates can be estimated from data [21] and adverse immunological effects can be assessed or excluded, our criterion may become highly valuable for decision-makers in countries facing vaccine shortages.

Although clinical studies of the approved SARS-CoV-2 vaccines may suggest that these vaccines are safe and effective, only little is known about their possible long-term adverse effects [51]. Clearly, for the comparison of different vaccination strategies we assume that negative long-term effects are negligible. In addition, we do not consider harm measures covering non-hospitalized symptomatic cases. Adverse effects and different levels of protection may be incorporated in models that account for different subgroups [52]. Yet, our results are independent of the actual fatality ratio for unvaccinated individuals.

To conclude, while current vaccine supplies are not keeping up with demand, especially in low- and middle-income countries, and newly-emerging variants of SARS-CoV-2 may reduce the effectiveness of currently available vaccines [53], it is desirable to provide decision makers with transparent tools that supports them in assessing different vaccination protocols. This study may be of help to healthcare officials and decision makers since, in contrast to existing literature, our combination of tools result in unexpectedly robust and highly decisive criteria. More generally, the presented framework establishes how epidemiologically efficient vaccine dosing strategies [54, 55] can be integrated into effective pandemic control plans.

## Data Availability

Our source codes are publicly available at https://github.com/lubo93/vaccination.

https://github.com/lubo93/vaccination

## ACKNOWLEDGEMENTS

LB acknowledges financial support from the SNF (P2EZP2_191888), NIH (R01HL146552), and Army Research Office (W911NF-18-1-0345). Parts of the simulations were performed on the ETH Euler cluster.

## DATA AND CODE AVAILABILITY

Our source codes are publicly available at https://github.com/lubo93/vaccination.

## MATERIALS AND METHODS

### Modeling prime and prime-boost vaccination

We adapt the SEIRD model [29, 32] to account for immunity waning and a vaccination-induced reduction in transmissibility [Fig. 1]. The fractions of susceptible, exposed, infected, recovered, and deceased individuals at time *t* are denoted by *S*(*t*), *E*(*t*), *I*(*t*), *R*(*t*), and *D*(*t*) respectively. Moreover, we denote the fractions of prime and prime-boost vaccinated susceptible individuals by *S*^★^(*t*) and *S*^★★^(*t*), respectively. With rate *ν*_1_, susceptible individuals get vaccinated with prime shots and with rate *ν*_2_ prime-vaccinated susceptible individuals get vaccinated with boost shots. The time dependence in the vaccination rates reflects temporal variations in the availability of vaccination doses, as explained below. The corresponding fractions of vaccinated exposed and infected individuals are denoted by *E*^★^(*t*) and *E*^★★^(*t*) and *I*^★^(*t*) and *I*^★★^(*t*). We use three constant rates *η*_1_, *η*_2_, *η*_3_ to model immunity waning (i.e., transitions from *S*^★^, *S*^★★^, and *R* to *S*). Characteristic time scales of waning immunity [20], defined by the inverse of the corresponding rates, are much longer than those associated with entering and leaving exposed and infected compartments, so we do not explicitly model waning immunity in these compartments. For long time horizons, additional birth and death processes may be employed to model birth and age-related death.

The resulting dynamics of the susceptible and exposed classes is described by the following rate equations:

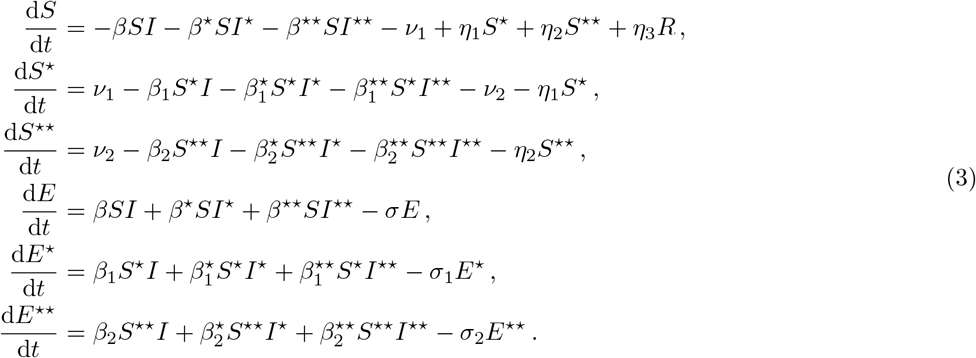

The maximum proportion of susceptible individuals that can be prime and prime-boost vaccinated is *S*(*t*) and *S*^★^(*t*), respectively. Based on vaccination data from Israel (Fig. 4), we assume linearly increasing immunization over time in our model and use the vaccination rates

**FIG. 4.**
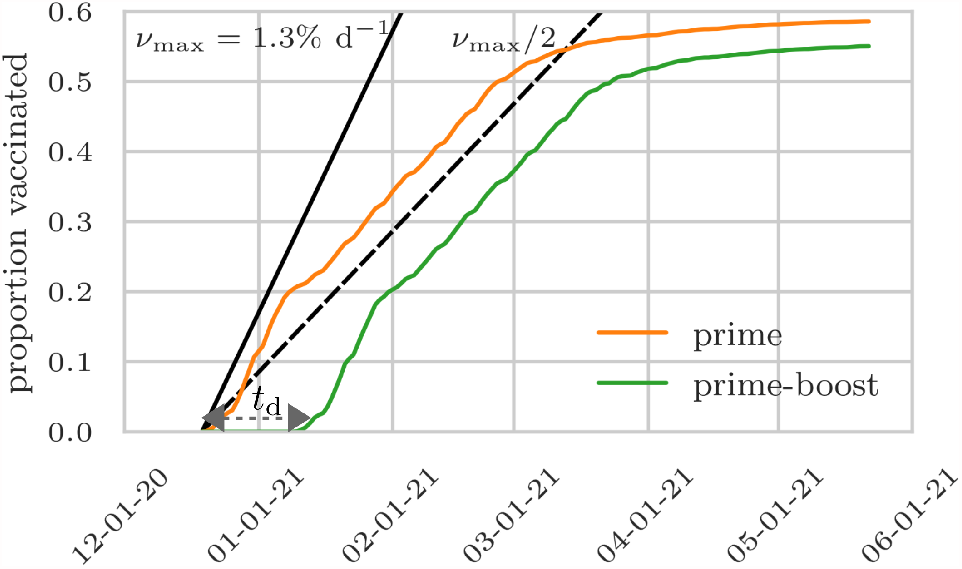
Proportions of prime and prime-boost vaccinated individuals in Israel. The proportions of prime (orange) and prime-boost (green) vaccinated individuals in Israel. The first prime shot was administered on December 20, 2020. On January 10, 2021 the first individuals received booster doses. The prime-boost delay *t*_*d*_ is thus 21 days. The solid and dashed black lines are guides to the eye, representing a maximum daily vaccination rate of *ν*_max_ = 1.3% and *ν*_max_ */*2, respectively. All data is taken from https://github.com/owid/covid-19-data/tree/master/public/data/vaccinations/country_data, accessed May 23, 2021.

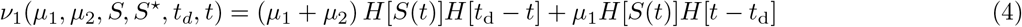

and

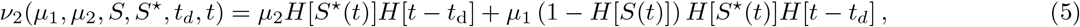

where *µ*_1_ = *ν*_max_ and *µ*_2_ = 0 for prime-first vaccination and *µ*_1_ = *µ*_2_ = *ν*_max_*/*2 for prime-boost vaccination. Here, *H*[*x*] denotes the Heaviside step function, which is zero for *x <* 0 and one for *x* ≥0. The function *H*[*t* −*t*_d_] describes the delay *t*_d_ of about 2–3 weeks [56] (Fig. 4) between prime and boost shots. Up to time *t*_*d*_, susceptible individuals get vaccinated with rate *µ*_1_ + *µ*_2_. If no susceptible individuals are left, prime-vaccinated individuals get vaccinated with rate *µ*_1_ too, leading to the term *µ*_1_(1 −*H*[*S*(*t*)])*H*[*S*^★^(*t*)]*H*[*t* −*t*_*d*_] in Eq. (5). In our model, only susceptible individuals are vaccinated. This can be justified by the assumption that susceptible individuals outnumber those in other disease states.

Exposed individuals transition to infected state at rates *σ, σ*_1_, and *σ*_2_. The evolution of the infected, recovered, and deceased compartments is described by:

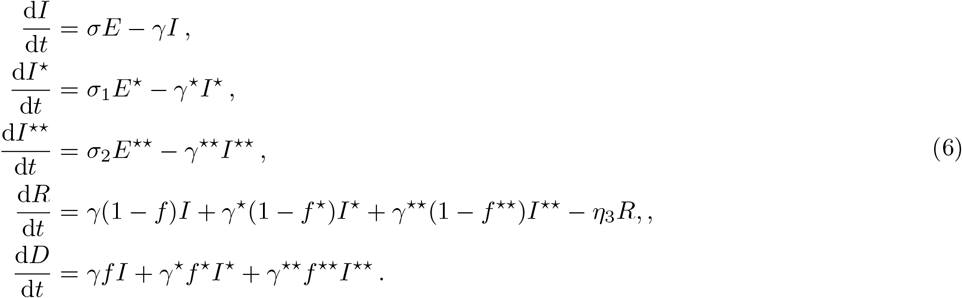

Only 10 of equations (3) and (6) are independent since we employ the normalization condition *S* + *S*^★^ + *S*^★★^ + *E* + *E*^★^ + *E*^★★^ + *I* + *I*^★^ + *I*^★★^ + *R* + *D* = 1. Different transmissibilities 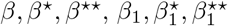, and 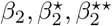 describe interactions between susceptible and infected individuals with different immunity levels.

For each infected compartment *I, I*^★^, and *I*^★★^, we calculate the infection fatality ratios (IFRs) [3] by dividing the associated cumulative number of deaths by the total number of infections in the unvaccinated, prime-vaccinated, and prime-boost-vaccinated compartments, respectively. The IFR of the unvaccinated pool of individuals is

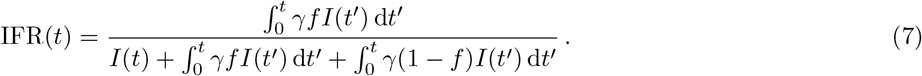

For constant *γ, f*, we obtain

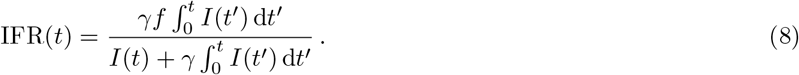

As the number of infected individuals approaches zero for long time horizons (i.e., lim_*t→∞*_ *I*(*t*) = 0), the IFR satisfies lim_*t→∞*_ IFR(*t*) = *f*. Similarly, lim_*t→∞*_ IFR^★^ (*t*) = *f*^★^ and lim_*t→∞*_ IFR^★★^ (*t*) = *f*^★★^ if *γ*^★^, *f*^★^ and *γ*^★★^, *f*^★★^ are time-independent.

Due to ergodicity breaking effects from multiplicative noise [57] deterministic models tend to overestimate infection and fatality. However, it is realistic to assume that the effects from noise are not discriminative as they do not differ for either vaccination campaign.

After all, the immunological intricacies of SARS-CoV-2 remain largely unknown and there is no single commonly accepted epidemiological standard model. At the same time, we anticipate more reliable data on immunity waning and other immunological effects in the coming months. Our framework is transparent and flexible enough to change or augment the (already high) degree of parameterization, or compartmentalization, if warranted.

### Basic reproduction number

We calculate the basic reproduction number *R*_0_ of the epidemic model (3) and (6) using the next-generation matrix method [61]. As a first step, we rewrite the rate equations (3) and (6) of the infected compartments in matrix form

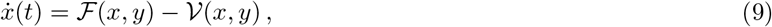

where *x* = (*E, E*^★^, *E*^★★^, *I, I*^★^, *I*^★★^)^*T*^, *y* = (*S, S*^★^, *S*^★★^, *R, D*), ℱ represents the vector of new infections, and 𝒱 describes all remaining transitions. We thus find for the corresponding Jacobians of ℱ and 𝒱 at the disease-free equilibrium:

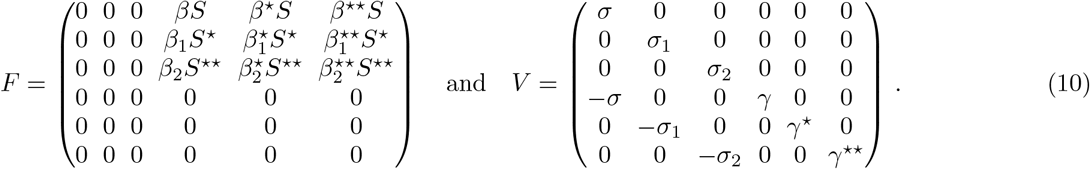

The basic reproduction number *R*_0_, the expected number of infections generated by an infectious individual in an otherwise completely susceptible population, is the spectral radius of the next-generation matrix *FV* ^−1^ [61]. Finding *R*_0_ for the general system (10) involves the analytically cumbersome task of finding roots of a cubic equation, which can be avoided by using numerical methods (e.g., the power method). For very effective vaccines, however, one may assume that the transmissibility of prime-boost vaccinated individuals is much lower than the transmissibility of unvaccinated individuals. That is, 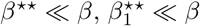, and 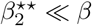. In this approximation, we obtain

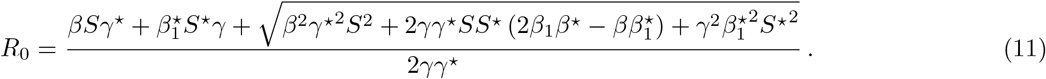

For *S*(0) = 1 and *S*^★^(0) = 0, the basic reproduction number is

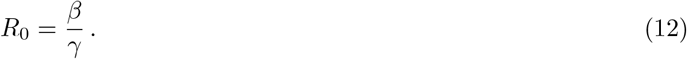

### Numerical solution and model parameters

To solve Eqs. (3) and (6) numerically, we use the Dormand–Prince method [62] with a maximum time step of 10^−1^ and simulate the evolution of different epidemics in the time interval [0, *T*] where *T* = 300 days. For the simulation results that we show in Fig. 2, we set *I*(0) = 10^−2^ and *S*(0) = 1 − *I*(0). If model parameters are held constant in Fig. 2, we use the parameters that are listed in Tab. I. Transmissibilities of infection events that involve at least one vaccinated individual are smaller than or equal to the baseline transmissibility *β* as long as mobility and distancing characteristics of vaccinated individuals do not differ significantly from those who are unvaccinated. In our model, this means that 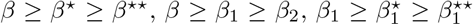, and 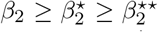 (equality holds for very ineffective vaccines). As vaccination campaigns and vaccine effectiveness analyses are ongoing, we used estimates for 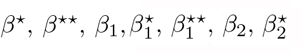 and 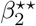 as reported in Tab. I. We also model the effect of small incidence rates and broader parameter ranges in a random-sampling analysis as reported in the next sections.

There are two more constraints that our model parameters have to satisfy to describe the impact of vaccination campaigns on disease transmission. First, the fatality ratio in the unvaccinated compartment is larger than the fatality ratios in the vaccinated compartments (i.e., *f* ≥ *f*^★^ ≥ *f*^★★^). We assume that differences in *f*^★^ and *f*^★★^ are negligible. Second, the waning rate in the prime-boost vaccinated compartment is smaller than the waning rate in the prime-vaccinated compartment (i.e., *η*_2_ ≤ *η*_1_).

### Influence of small incidence rates

To study the effect of small incidence rates on the location and extent of prime-first and prime-boost preference regions, we set *I*(0) = 10^−5^ and 10^−7^, which is three to five orders of magnitude smaller than the value *I*(0) = 10^−2^ we used in Fig. 2, and show vaccination preference diagrams for *η*_1_ − *η*_2_ vs. *ν*_max_ and *η*_1_ − *η*_2_ vs. *R*_0_ in Fig. 5. We observe that a threshold *η*_1_ − *η*_2_ = 0.01 separates prime-first and prime-boost preference regions in both diagrams. This value is smaller than the threshold of *η*_1_ − *η*_2_ = 0.017, which we used in Fig. 2.

**FIG. 5.**
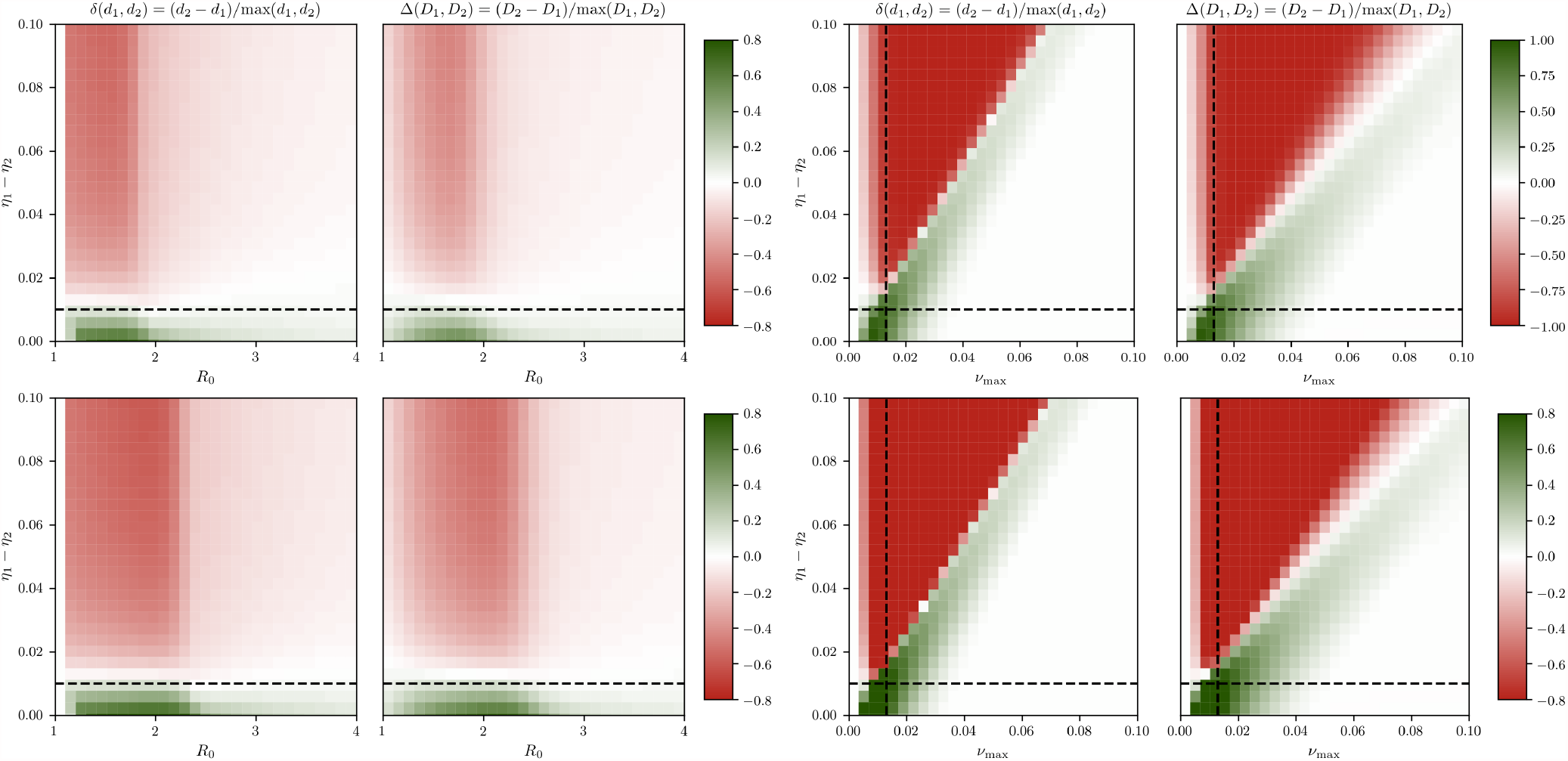
Selection of vaccination-campaign-preference diagrams for small incidence rates. For selected combinations of basic reproduction number *R*_0_, waning rate difference *η*_1_ − *η*_2_, and maximum vaccination rate *ν*_max_, we plot RFC-*δ* [Eq. (1)] and RFC-Δ [Eq. (2)]. Green-shaded regions indicate preference for prime (RFC-*δ* > 0, RFC-Δ > 0), red-shaded regions indicate preference for prime-boost (RFC-*δ* < 0, RFC-Δ < 0). In the top panels, we set *I*(0) = 10^−5^; in the bottom panels, we set *I*(0) = 10^−7^. The remaining parameters are as in Tab. I. Dashed lines are guides to the eye: Thresholds *η*_1_ − *η*_2_ = 0.01, and Israel’s vaccination rate as of Feb. 1, 2021 [4] (*ν*_max_ = 0.013).

### Empirical vaccination data

We also study the influence of empirical vaccination data (Fig. 4) on the prime-first preference threshold *η*_1_ − *η*_2_ = 0.017. Figure 6 shows that preference for prime-first over prime-boost vaccination is given for faster immunity waning than indicated by the threshold *η*_1_ − *η*_2_ = 0.017, demonstrating the robustness of the original waning-rate difference threshold.

**FIG. 6.**
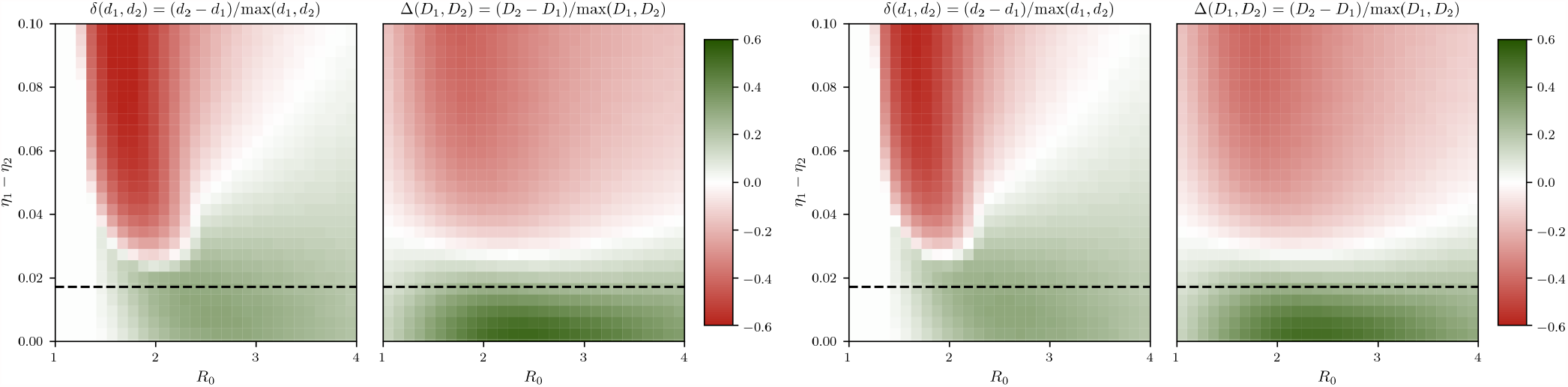
Empirical vaccination data from Israel does not lower decision threshold, *η*_1_ − *η*_2_. For variations in *η*_1_ − *η*_2_ and *R*_0_, we plot RFC-*δ* [Eq. (1)] and RFC-Δ [Eq. (2)] for empirical vaccination data (Fig. 4) and two natural immunity waning rates. In the left panel, we set *η*_3_ = 6 ×10^−3^*/*day (waning within about 6 months); in the right panel, we set *η*_3_ = 3 ×10^−3^*/*day (waning within about 12 months). Green-shaded regions indicate preference for prime (RFC-*δ >* 0, RFC-Δ > 0), red-shaded regions indicate preference for prime-boost (RFC-*δ <* 0, RFC-Δ < 0). The simulation horizon is *T* = 150 days. The remaining parameters are as in Tab. I. Dashed lines are guides to the eye of the original threshold *η*_1_ − *η*_2_ = 0.017. We observe preference for prime-first over prime-boost vaccination even for faster waning, *η*_1_ − *η*_2_ > 0.017, compared to other epidemiological scenarios with a constant and, on average, larger vaccination rate.

### Influence of natural immunity waning

For the critical time horizon of a few months that we consider in the main text, for scenarios (referred to as datasets) A and B, we assume robust natural (T cell) immunity in accordance with corresponding clinical data [63]. Figure 7 shows vaccination-campaign-preference diagrams for *η*_1_ − *η*_2_ vs. *R*_0_, and for natural immunity waning time scales of about 6 and 12 months. We observe that these variations in *η*_3_ do not change the campaign-preference threshold (dashed black line in Fig. 7).

**FIG. 7.**
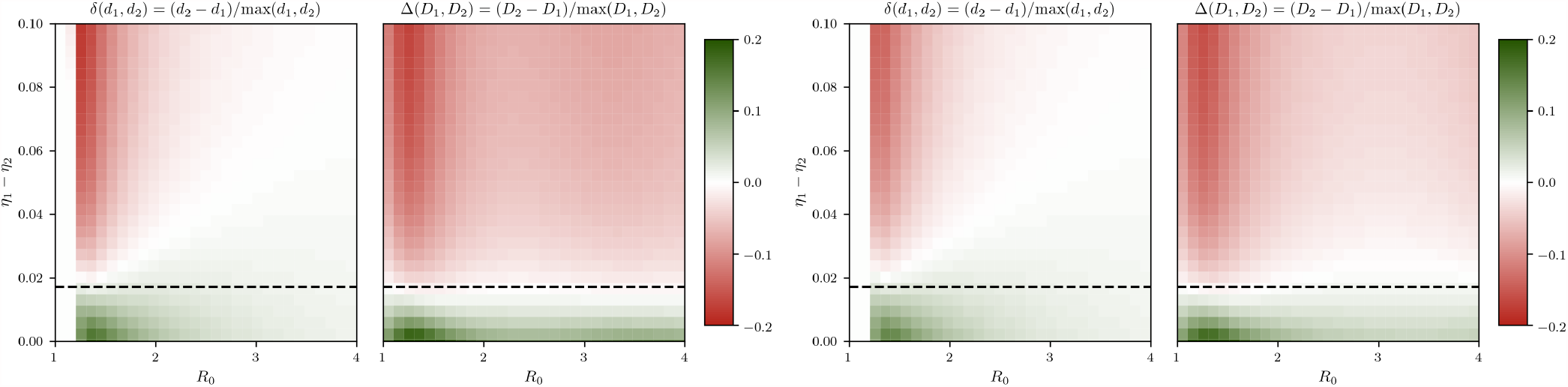
Natural immunity waning does not lower decision threshold, *η*_1_ − *η*_2_. For variations in *η*_1_ − *η*_2_ and *R*_0_, we plot RFC-*δ* [Eq. (1)] and RFC-Δ [Eq. (2)] for two natural immunity waning rates. In the left panel, we set *η*_3_ = 6 ×10^−3^*/*day (waning within about 6 months); in the right panel, we set *η*_3_ = 3 ×10^−3^*/*day (waning within about 12 months). Green-shaded regions indicate preference for prime (RFC-*δ* > 0, RFC-Δ > 0), red-shaded regions indicate preference for prime-boost (RFC-*δ* < 0, RFC-Δ < 0). The remaining parameters are as in Tab. I. Dashed lines are guides to the eye of the original, unchanged threshold *η*_1_ − *η*_2_ = 0.017.

### Monte Carlo sampling

The parameter distributions that we use in our random sampling and decision tree analysis are summarized in Tab. II. We generate two datasets with *N* = 50000 samples each and analyze the influence of different combinations of model parameters and initial conditions on RFC-*δ*(*d*_1_, *d*_2_) [Eq. (1)] and RFC-Δ(*D*_1_, *D*_2_) [Eq. (2)].

### Correlation between fatality measures

RFC-*δ* and RFC-Δ are complementary fatality measures but are correlated, [Fig. 8 (A–C), *R* = 0.94 (A), *R* = 0.68 (B), and *R* = 0.68 (C); corresponding p-values are smaller than machine precision]. The correlation observed for the threshold combination *ν*_max_ ≤0.013 day^−1^ and *η*_1_ − *η*_2_ ≤0.017 day^−1^ confirms the discriminative power and robustness of our results regarding the choice of both fatality measures.

**FIG. 8.**
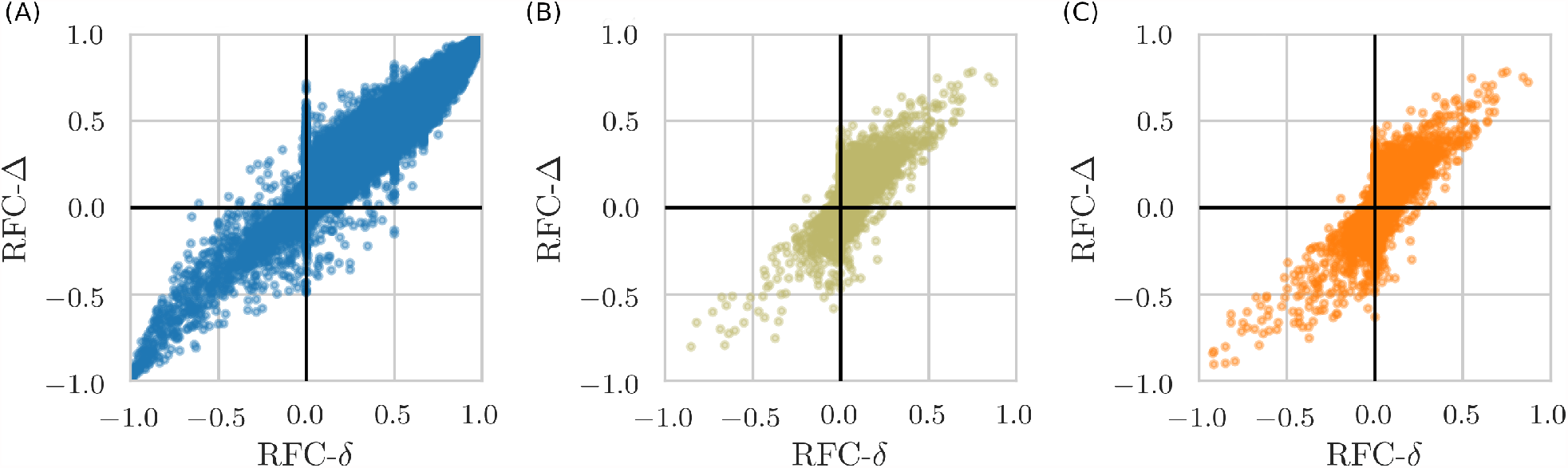
Fatality measure correlations for entire and restricted high-dimensional parameter spaces. Correlation plots of RFC-*δ* and RFC-Δ. Unconditioned data (orange) is compared with two conditioned, (i) data shown in beige: *ν*_max_ ≤0.047 day^−1^ and *η*_1_ − *η*_2_ ≤0.056 day^−1^, and (ii) data shown in blue: *ν*_max_ ≤0.013 day^−1^ and *η*_1_ − *η*_2_ ≤ 0.017 day^−1^. Thresholds (i) are inferred from our decision tree analysis. The presented data are based on 5 ×10^4^ (blue and orange curves) and about 4.4 ×10^4^ (beige curves) samples of the entire 25-dimensional parameter space. Parameters are as listed in Tab. II (no natural immunity waning).

### Influence of natural immunity waning

We further analyze the effect of natural immunity waning by sampling *η*_3_ from 𝒰(0, 0.1) day^−1^. Figure 9 shows the corresponding distributions and correlation plots associated with the fatality measures RFC-*δ* and RFC-Δ.

**FIG. 9.**
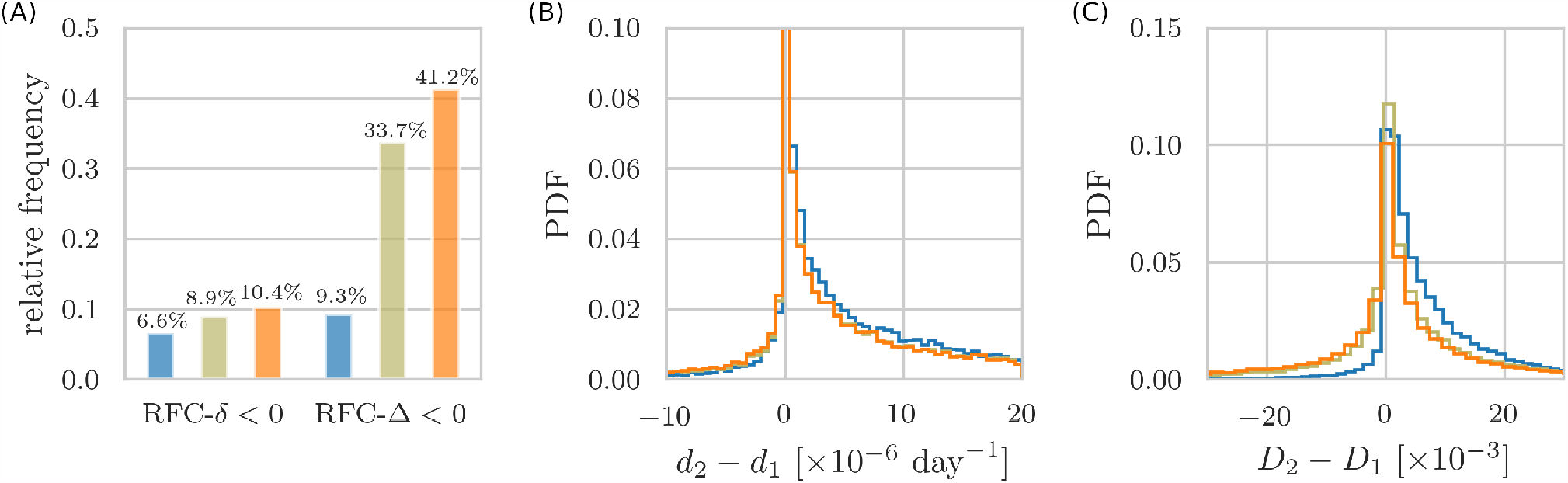
Monte Carlo sampling of entire and restricted high-dimensional parameter spaces with natural immunity waning. Unconditioned data (orange) is compared with two conditioned, (i) data shown in beige: *ν*_max_ ≤0.047 day^−1^ and *η*_1_ − *η*_2_ ≤0.056 day^−1^, and (ii) data shown in blue: *ν*_max_ ≤0.013 day^−1^ and *η*_1_ − *η*_2_ ≤ 0.017 day^−1^. Thresholds (i) are inferred from our decision tree analysis. We sampled natural immunity waning rates *η*_3_ from the distribution 𝒰 (0, 0.1) day^−1^. All remaining parameters are specified in Tab. II. (A) Relative frequency of prime-boost-preference samples (RFC-*δ <* 0, RFC-Δ < 0) for the three datasets (similar to Fig. 3(A) of the main text). Error bars are below 0.5% (not shown). (B) Probability density function (PDF) of the difference between death rates *d*_2_ (prime-boost) and *d*_1_ (prime). For the conditioned data, averages of *d*_1_ − *d*_2_ are about 1.0 ×10^−6^ day^−1^ (blue curve) and about 8.3 ×10^−7^ day^−1^ (beige curve), larger than the mean 6.8 ×10^−7^ day^−1^ of the unconditioned data. (C) PDF of the difference between the total number of deaths *D*_2_ (prime-boost) and *D*_1_ (prime). For the conditioned data, the means of *D*_1_ − *D*_2_ are about 7.3 × 10^−3^ (blue curve) and about 2.0 × 10^−3^ (beige curve), larger than the mean −1.5 × 10^−3^ of the unconditioned data.

The relative frequencies of prime-boost-preference samples, characterized by RFC-*δ <* 0 and RFC-Δ < 0, are estimated as 10.4% (SE: 0.3%) and 41.2% (SE: 0.4%), see orange bars in Fig. 9(a). For waning rate differences *η*_1_ − *η*_2_ ≤0.056 day^−1^ and vaccination rates *ν*_max_ ≤ 0.047 day^−1^, the proportions of prime-boost-preference samples are 8.9% (SE: 0.3%) for RFC-*δ <* 0 and 33.7% (SE: 0.4%) for RFC-Δ < 0 [beige bars in Fig. 9(a)]. Using the condition *η*_1_ − *η*_2_ < 0.017 day^−1^ (dashed black lines in Fig. 2) and vaccination rates *ν*_max_ < 0.013 day^−1^ [4] leads to proportions of prime-boost-preference samples of 6.6% (SE: 0.2%) for RFC-*δ <* 0 and 9.3% (SE: 0.3%) for RFC-Δ < 0 [blue bars in Fig. 9(a)].

As in the main text, we find that constraining the studied parameter space by lowering *ν*_max_ and *η*_1_ − *η*_2_ yields an substantial increase in prime-first-preference samples that are associated with fewer total fatalities, as quantified by RFC-Δ. The proportions of prime-boost preference samples with RFC-*δ <* 0 fall into the narrow rage between 7 and 10% and are less affected by the chosen parameter restrictions [Fig. 9(a)]. This again supports the robustness of our results. For randomly sampled parameters that account for different natural immunity-waning rates, prime-first vaccination is preferred regarding RFC-*δ*.

### Risk-group stratification

To study the effect age-related fatality rates, we perform a random-sampling analysis for two age groups. In the first group, we set *f* ∼ 𝒰 (10^−4^, 10^−3^), *f* ^∗^ ∼ 𝒰 (10^−4^, *f*), *f* ^∗^ = *f*^∗ ∗^. Fatality rates *f* of less than 0.1% have been observed for individuals younger than 40 years [60]. In the second group, we set *f* ∼ 𝒰 (10^−3^, 10^−2^), *f* ^∗^ ∼ 𝒰 (10^−3^, *f*), *f* ^∗^ = *f* ^∗ ∗^. Fatality rates of about 0.1–1% have been reported for individuals with an age between 40–70 years. As in the previous section, we also account for natural immunity waning by setting *η*_3_ ∼ 𝒰 (0, 0.1) day^−1^. All remaining parameters are specified in Tab. II.

Figure 10 shows different distributions and correlation plots associated with the fatality measures RFC-*δ* and RFC-Δ for both age groups. The shown results are in agreement with those reported in the previous section and main text. Prime-boost-preference samples (i.e., those samples with RFC-*δ <* 0 and RFC-Δ < 0) occur less frequently than prime-first-preference samples in both age groups. The conditions *η*_1_ − *η*_2_ < 0.017 day^−1^ (dashed black lines in Fig. 2) and *ν*_max_ < 0.013 day^−1^ [4] again lead to significantly reduced proportions of prime-boost-preference samples (blue bars and curves in Fig. 10), supporting the validity of these decisive thresholds.

**FIG. 10.**
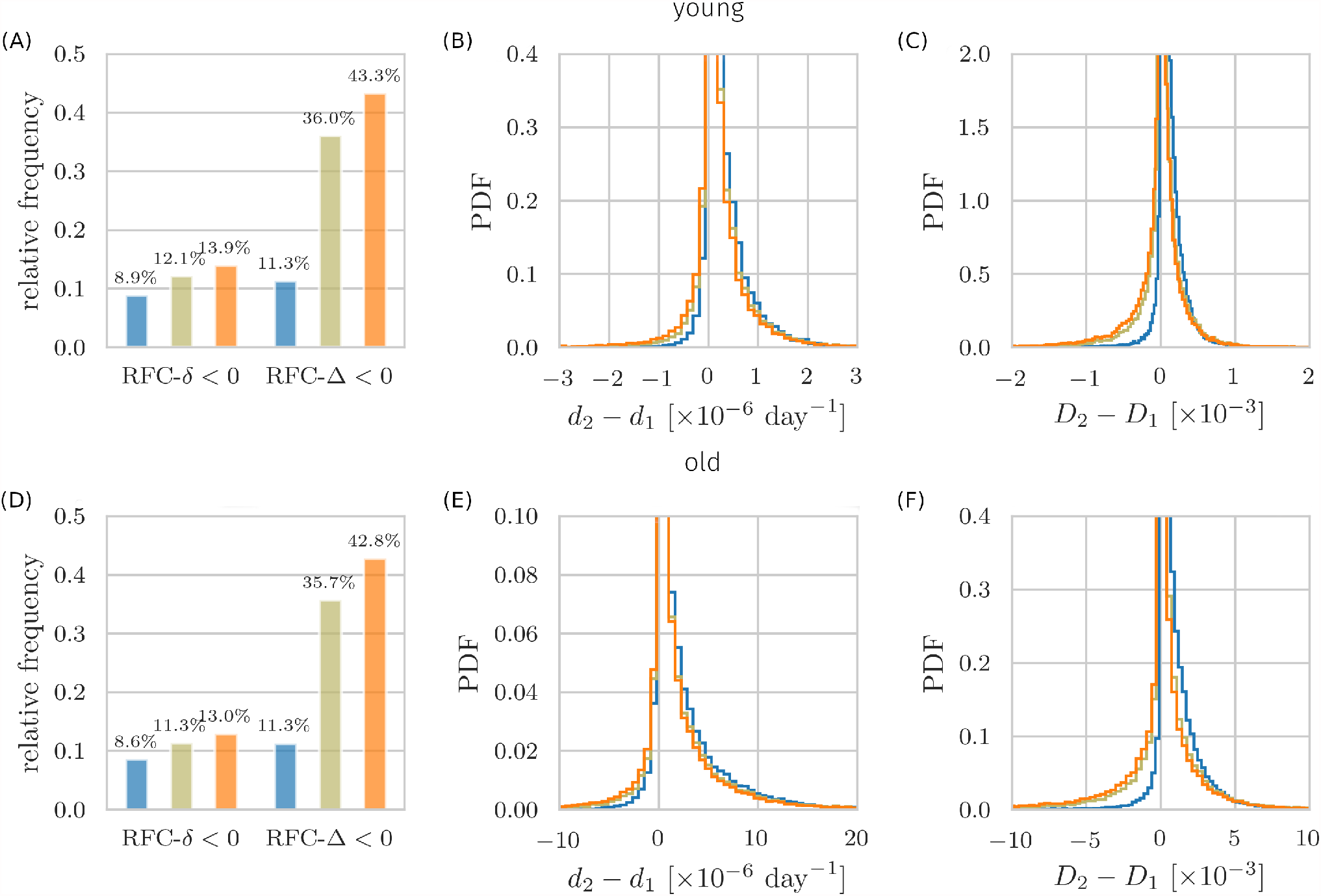
Monte Carlo sampling of entire and restricted high-dimensional parameter spaces with age-stratification and natural immunity waning. Unconditioned data (orange) is compared with two conditioned, (i) data shown in beige: *ν*_max_ ≤ 0.047 day^−1^ and *η*_1_ − *η*_2_ ≤ 0.056 day^−1^, and (ii) data shown in blue: *ν*_max_ ≤ 0.013 day^−1^ and *η*_1_ − *η*_2_ ≤ 0.017 day^−1^. Thresholds (i) are inferred from our decision tree analysis. (A–C) Fatality rates are *f* ∼ 𝒰 (10^−4^, 10^−3^), *f* ^∗^ ∼ 𝒰 (10^−4^, *f*), and *f* ^∗^ = *f* ^∗ ∗^. (D–F) Fatality rates are *f* ∼ 𝒰 (10^−3^, 10^−2^), *f* ^∗^ ∼ 𝒰 (10^−3^, *f*), and *f* ^∗^ = *f* ^∗ ∗^. Natural immunity waning rate *η*_3_ is sampled from 𝒰 (0, 0.1) day^−1^. All remaining parameters are specified in Tab. II. (A,D) Same plots as in Fig. 3 of the main text. Relative frequency of prime-boost-preference samples (RFC-*δ <* 0, RFC-Δ < 0) for the three datasets. Error bars are below 0.5% (not shown). (B,E) Probability density function (PDF) of the difference between death rates *d*_2_ (prime-boost) and *d*_1_ (prime). For the conditioned data in (B), averages of *d*_1_ − *d*_2_ are about 1.2 ×10^−8^ day^−1^ (blue curve) and about 8.3 ×10^−9^ day^−1^ (beige curve), larger than the mean 6.1 ×10^−9^ day^−1^ of the unconditioned data. For the conditioned data in (E), averages of *d*_1_ −*d*_2_ are about 1.1 ×10^−7^ day^−1^ (blue curve) and about 8.7 ×10^−8^ day^−1^ (beige curve), larger than the mean 6.6 ×10^−8^ day^−1^ of the unconditioned data. (C,F) PDF of the difference between the total number of deaths *D*_2_ (prime-boost) and *D*_1_ (prime). For the conditioned data in (C), the means of *D*_1_ − *D*_2_ are about 9.1 ×10^−5^ (blue curve) and about 1.6 × 10^−5^ (beige curve), larger than the mean 3.2 × 10^−5^ of the unconditioned data. For the conditioned data in (F), the means of *D*_1_ − *D*_2_ are about 8.9 ×10^−4^ (blue curve) and about 1.7 ×10^−4^ (beige curve), larger than the mean −2.7 ×10^−4^ of the unconditioned data.

### Decision Tree Analysis

A binary decision tree consists of a root condition and branches, where the left branch refers to the “yes”-branch while the rights branch refers to the “no”-branch.

We employed binary decision tree learning with repeated stratified cross validation (*k* = 5 folds, *n* = 10 repeats). The algorithm `RepeatedStratifiedKFold` (available in the Python library scikit-learn^1^) optimizes for split purity using Gini as loss function (split criterion).

Gini impurity is a standard measure in tree learning that quantifies how often a randomly chosen sample from the training dataset would be incorrectly labeled if it was entirely randomly labeled, given the distribution of (binary) labels in the subset. In our analysis, labels are “prime-first” and “prime-boost”.

Stratified cross-validation is based on splitting the data into folds such that each fold has the same proportion of observations with a given categorical value. It is particularly useful for imbalanced datasets. Overall, here, we have more prime-first samples than prime-boost ones.

Following standard procedure, we split the dataset (randomly) into **training** and **test** datasets, 70% and 30%, respectively. Learning is performed using the training dataset while the test dataset is cross-validated.

The training accuracy score (for a binary classification task) is defined as the relative number of correctly predicted labels, that is,

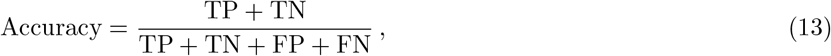

where TP are true positives, TN are true negatives, FP are false positives, and FN are false negatives.

In our binary classification problem, positives are prime-first labeled samples, negatives are prime-boost labeled samples. Class prime-boost is defined by Δ*D <* 0 (red-shaded regions in Fig. 2). Class prime-first is defined by Δ*D >* 0 (green-shaded regions in Fig. 2).

An *n*-times repeated stratified cross-validation is based on the following iteration: (i) Shuffle the test dataset randomly, (ii) split the dataset into *k* folds, (iii) for each fold: take the fold as test dataset and take the remaining folds as training dataset, (iv) fit the tree on the training dataset and evaluate it on the test dataset.

Accuracy and balanced accuracy are monitored as main cross-validation scores. We also monitored precision, F1-score based metrics, ROC AUC, and recall. Balanced accuracy is warranted for imbalanced datasets and defined as the arithmetic mean of 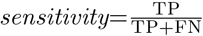 (true positive rate) and 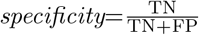 (true negative rate).

*Dataset A* Dataset A comprises of 50000 randomly sampled data points for parameter ranges and disease stages as described in Tab. II (without natural immunity waning). In Fig. 3 dataset A is called “unconditioned data” (displayed in orange).

Here, we analyze dataset A, see Fig. 11. Training performance is excellent and reaches 100% for large depths due to overfitting. Learning performance is satisfactory, as seen from similar behaviors for test accuracy and balanced accuracy, around 70% for depth = 3. The resulting tree reveals two highly discriminative conditions for prime-boost preference, *ν*_max_ ≤ 0.047 and *η*_1_ − *η*_2_ ≤0.056. This threshold combination is used in Fig. 3, referred to as the conditioned data, displayed in beige.

**FIG. 11.**
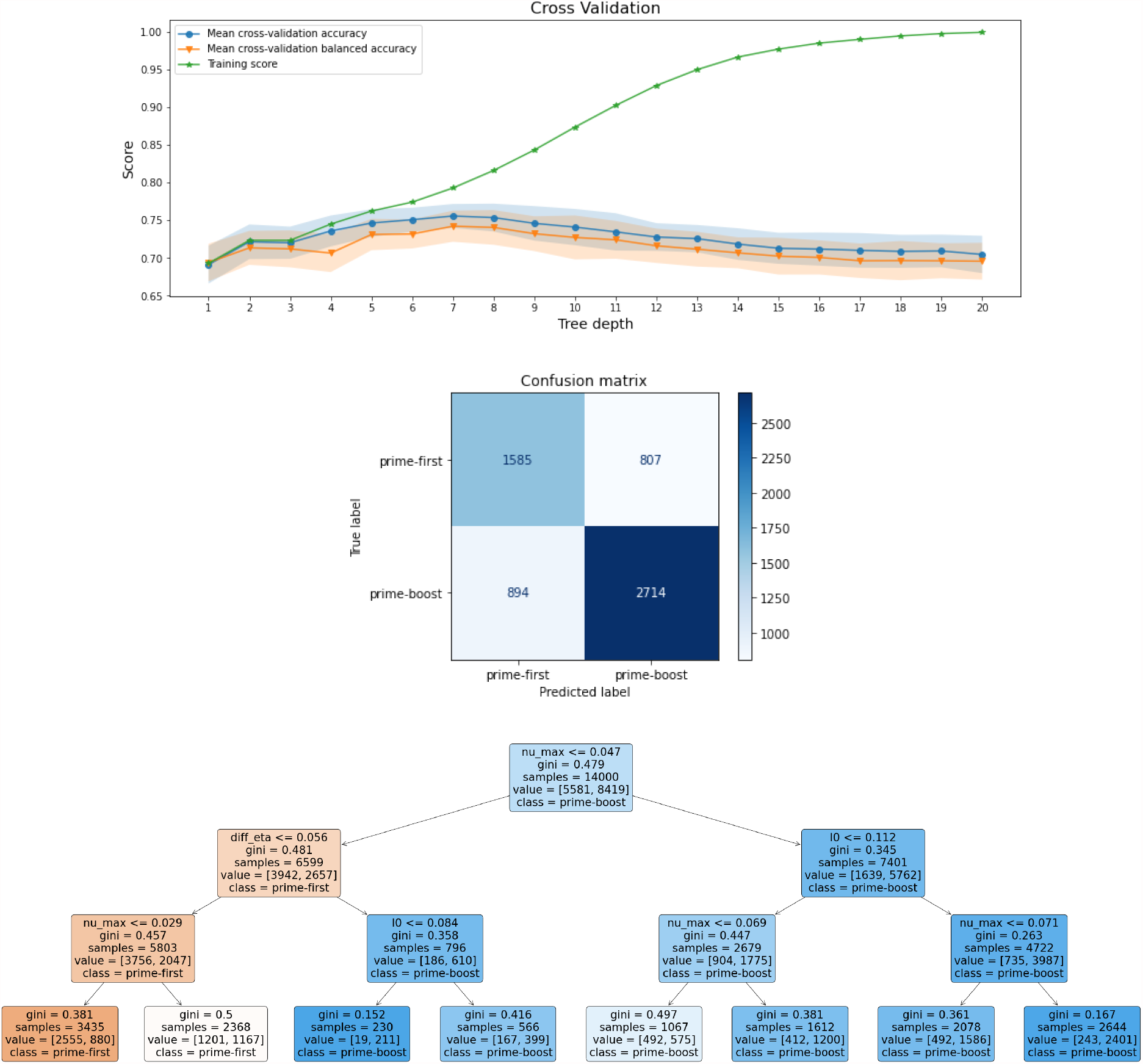
Decision tree analysis for dataset A. Cross-validation: Training accuracy (green curve) reaches 100% for large tree size. This indicates that fitting is satisfactory, while test training data is overfitted for large tree sizes, as expected. Accuracy (blue curve and shades) and balanced accuracy (orange curve and shades) show a fair and also similar performance for all depths. This indicates satisfactory class prediction. Confusion matrix: Number of instances for predicted and true (ground truth) labels, for prime-first and prime-boost. Upper left: True positives, Upper right: False positives, Lower left: False negatives, Lower right: True negatives. Notation: Positives are prime-first, negatives prime-boost. Decision tree: (*Value*) denotes the number of samples in class “prime-first” (left part) and class “prime-boost” (right part), respectively, for the given branch (brackets), while *samples* denote the total sum of samples at the given branch. Left branches satisfy the displayed condition (“yes” branch), right descendants are “no” branches. Notation: diff_eta = *η*_1_ − *η*_2_, nu_max = *ν*_max_, and *I*0 = *I*(0). Shown tree: depth = 3.

*Dataset B* Here we study the conditioned data constrained by *η*_1_ − *η*_2_ ≤ 0.017 and *ν*_max_ < 0.013, as analyzed in Fig. 3 (blue), here called dataset B. For this dataset, we uniformly sampled initial proportions of infected individuals, *I*(0), on a logarithmic scale from 10^−7^ to 3 × 10^−1^. This way of sampling allows us to study the robustness of decision boundary thresholds for a large range of initial disease prevalences.

Dataset B comprises of 50000 randomly sampled data points (without natural immunity waning) where samples simultaneously satisfy *η*_1_ − *η*_2_ ≤0.017 and *ν*_max_ < 0.013. Class prime-boost is defined by Δ*D <* 0 (red-shaded regions in Fig. 2). Class prime-first is defined by Δ*D >* 0 (green-shaded regions in Fig. 2).

Results are presented in Fig. 12. The training accuracy curve (green curve) shows high accuracy levels from overfitting of the **training** set that reaches 100% for large tree depths. The accuracy curve (blue) shows the mean of the cross-validation of the accuracy for the **test** dataset.

**FIG. 12.**
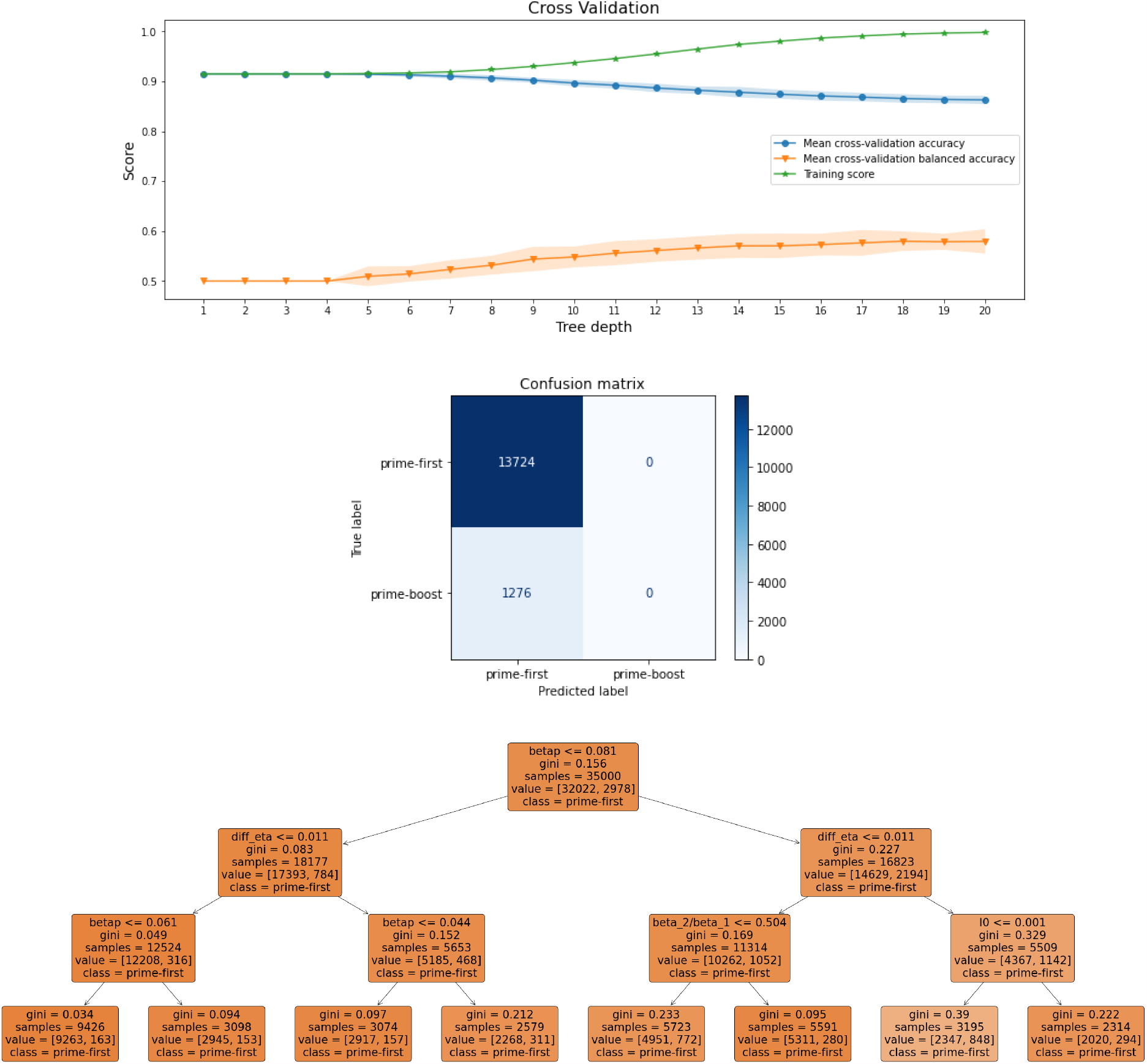
Decision tree analysis for dataset B. Cross-validation: Large spread between training accuracy (green curve) and balanced accuracy (orange) for all depths indicate poor learning performance regarding a possible subtree structure of prime-boost samples but excellent class prediction. High values of accuracy results from excellent prediction for prime-first samples (majority 13724) while prediction of prime-boost is poor (1276 false negatives, 0 true negatives in confusion matrix). Confusion matrix: Number of instances for predicted and true (ground truth) labels, for prime-first and prime-boost. Upper left: True positives, Upper right: False positives, Lower left: False negatives, Lower right: True negatives. Notation: Positives are prime-first, negatives prime-boost. Decision tree: (*Value*) denotes the number of samples in class “prime-first” (left part) and class “prime-boost” (right part), respectively, for the given branch (brackets), while *samples* denote the total sum of samples at the given branch. Left branches satisfy the displayed condition (“yes” branch), right descendants are “no” branches. Notation: betap = *β*^★^, diff_eta = *η*_1_ − *η*_2_, beta_2*/*beta_1 = *β*_2_ */β*_1_, *I*0 = *I*(0). Shown depth = 3. Resulting tree of depth = 3 is essentially equivalent to *always prime-first*, leading to an accuracy of 92%. With increasing depth balanced accuracy (and recall) increase only slightly.

The results confirm that no conditions other than the constraints *η*_1_ − *η*_2_ ≤ 0.017 and *ν*_max_ < 0.013 robustly characterize prime-first preference domains.

## Footnotes

1 https://scikit-learn.org/stable/modules/generated/sklearn.model_selection.RepeatedStratifiedKFold.html, accessed: 02-28-2021

